# HORNET: Tools to find genes with causal evidence and their regulatory networks using eQTLs

**DOI:** 10.1101/2024.10.28.24316273

**Authors:** Noah Lorincz-Comi, Yihe Yang, Jayakrishnan Ajayakumar, Makaela Mews, Valentina Bermudez, William Bush, Xiaofeng Zhu

## Abstract

**Motivation:** Nearly two decades of genome-wide association studies (GWAS) have identify thousands of disease-associated genetic variants, but very few genes with evidence of causality. Recent methodological advances demonstrate that Mendelian Randomization (MR) using expression quantitative loci (eQTLs) as instrumental variables can detect potential causal genes. However, existing MR approaches are not well suited to handle the complexity of eQTL GWAS data structure and so they are subject to bias, inflation, and incorrect inference.

**Results:** We present a whole-genome regulatory network analysis tool (HORNET), which is a comprehensive set of statistical and computational tools to perform genome-wide searches for causal genes using summary level GWAS data that is robust to biases from multiple sources. Applying HORNET to schizophrenia, we identified differential magnitudes of gene expression causality. Applying HORNET to schizophrenia, we identified differential magnitudes of gene expression causality across different brain tissues.

**Availability and Implementation:** Freely available at https://github.com/noahlorinczcomi/HORNETor Mac, Windows, and Linux users.

## 1 Introduction

Genetic epidemiologists have spent decades trying to identify genes that cause disease [26]. Significant effort has been given to experimental methods [42, 49], linkage studies [39], genome-wide association studies (GWAS), and functional annotation of putative disease-associated genetic variants [48]. These methods of causal validation may be costly, may not always provide causal inference, and have sometimes produced conflicting results [31]. They also generally cannot be scaled to efficiently test hundreds or thousands of genes simultaneously. Cis Mendelian Randomization (*cis*MR) has been proposed as a cost-and time-efficient alternative to identify potential causal genes and can leverage the wealth of publicly available summary data from genome-wide association studies (GWAS) and eQTL studies [22, 40, 51, 60]. In this context, cis MR uses instrumental variables that are gene expression quantitative trait loci (eQTLs) to estimate tissue-specific causal effects of gene expression on disease risk [19]. Cis MR methods are similar to transcriptome-wide association study (TWAS) methods, which test the association between predicted gene expression and the outcome phenotype. TWAS may suffer from reduced power due to imprecise estimation of gene expression in the discovery population [12, 32, 52], and from direct SNP associations with the outcome phenotype, known as horizontal pleiotropy. MR requires only GWAS summary statistics and a range of robust tools to control the Type I error and bias from horizontal pleiotropy rate have been developed [28, 34]. The MR-based approach can either consider each gene separately (univariable MR) or jointly with surrounding genes in a regulatory network (multivariable MR). Since it is well known that many genes are members of large regulatory networks [16, 29], multivariable MR may be better suited to study multiple gene expressions simultaneously than univariable MR that study one gene expression and one trait separately, such as TWAS [33, 34, 44].

However, there is currently no unified statistical or computational framework for applying multivariable MR to the study of causal genes. Performing multivariable MR with summary data from eQTL and disease GWAS (eQTL-MVMR) has many challenges, including the handling of missing data, linkage disequilibrium (LD) between eQTLs, gene tissue specification, gene prioritization, and causal inference. Without careful attention to each of these challenges, the simple application of traditional multivariable MR methods to these data may produce spurious results which may fail in follow-up experimental testing. We present HORNET, a set of bioinformatic tools that can be used to robustly perform eQTL-MVMR with GWAS summary data. We demonstrate that existing univariable and multivariable implementations of eQTL-MR are vulnerable to biases and/or inflated Type I and II error rates from weak eQTLs, correlated horizontal pleiotropy (CHP), high correlations between genes, missing data, and misspecified LD structure.

## 2 System and Methods

### 2.1 Data

HORNET uses summary level data from GWAS of cis gene expression (eQTL) and a disease phenotype. cis-eQTL GWAS data should generally provide estimates of association between the expression of each gene and all SNPs within ±1Mb of them. These data are publicly available from consortia such as eQTLGen [54] and the Genotype-Tissue Expression (GTEx) project [10]. Disease GWAS data can typically be downloaded from public repositories such as the GWAS Catalog [46]. HORNET additionally requires an LD reference panel with corresponding `.bim, .bed`, and `.fam files`. The 1000 Genomes Phase 3 (1kg) [9] reference panel is automatically included with the HORNET software for African, East Asian, South Asian, European, Hispanic, and trans-ancestry populations, although researches may use their own reference panels such as those from the UK Biobank [47].

### 2.2 Instrument selection and missing data

Selection of the IV set in eQTL-MVMR using standard IV selection methods can either reduce statistical power or make estimation of causal effects impossible because of the structure of cis-eQTL GWAS summary statistics. Univariable eQTL-MR for the *k*th gene in a locus of *p* genes uses the set 𝒮_*k*_ of cis-eQTLs as IVs and performs univariable regression [21]. Multivariable eQTL-MR in the same locus uses the superset 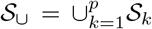 and performs multivariable regression [40]. Since most publicly available cis-eQTL data only contain estimates of association between SNPs and all genes within ±1Mb of them (e.g., [10, 54]), not all SNPs in 𝒮_∪_ may have association estimates that are present in the data. An alternative approach is to use the set 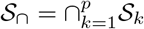 which contains SNPs with association estimates that are available for all *p* genes. However, this set may contain very few SNPs, if any, for some relatively large loci which contain many genes that are co-regulated. If the size of 𝒮_∩_ is small, there can be limited statistical power for eQTL-MVMR because the power in MR is proportional to the total trait variance explained by the IVs [34]. Thus, only 𝒮_∪_ is used in HORNET.

We propose imputing missing data using one of three approaches that users of HORNET can choose between: (i) imputation of missing values with 0s, (ii) imputation based only on LD structure between observed and unobserved SNPs [43], and (iii) imputation based on a modified matrix completion algorithm (MV-Imp). Using any of these methods, only estimates of association between SNPs and the gene expression phenotype are imputed. The MV-Imp approach in (iii) is applied to SNPs in the union set 𝒮_∪_ and presented in Algorithm 1. This approach assumes a low-rank structure of the MR design matrix and accounts for estimation error and LD structure. As mentioned, public cis-eQTL summary data are generally available for SNP-gene pairs within ±1Mb of each other. Using individual-level data from 236 unrelated non-Hispanic White subjects, we demonstrate in Figure 4 of the **Supplement** that association estimates outside of the 1Mb window have mean 0 and constant variance with high probability. Imputation using MV-Imp imputes data with the lowest error in simulation 2, though imputation of missing values with zeros performs similarly and is more computationally efficient.

#### Algorithm 1

Pseudo-code of eQTL imputation.

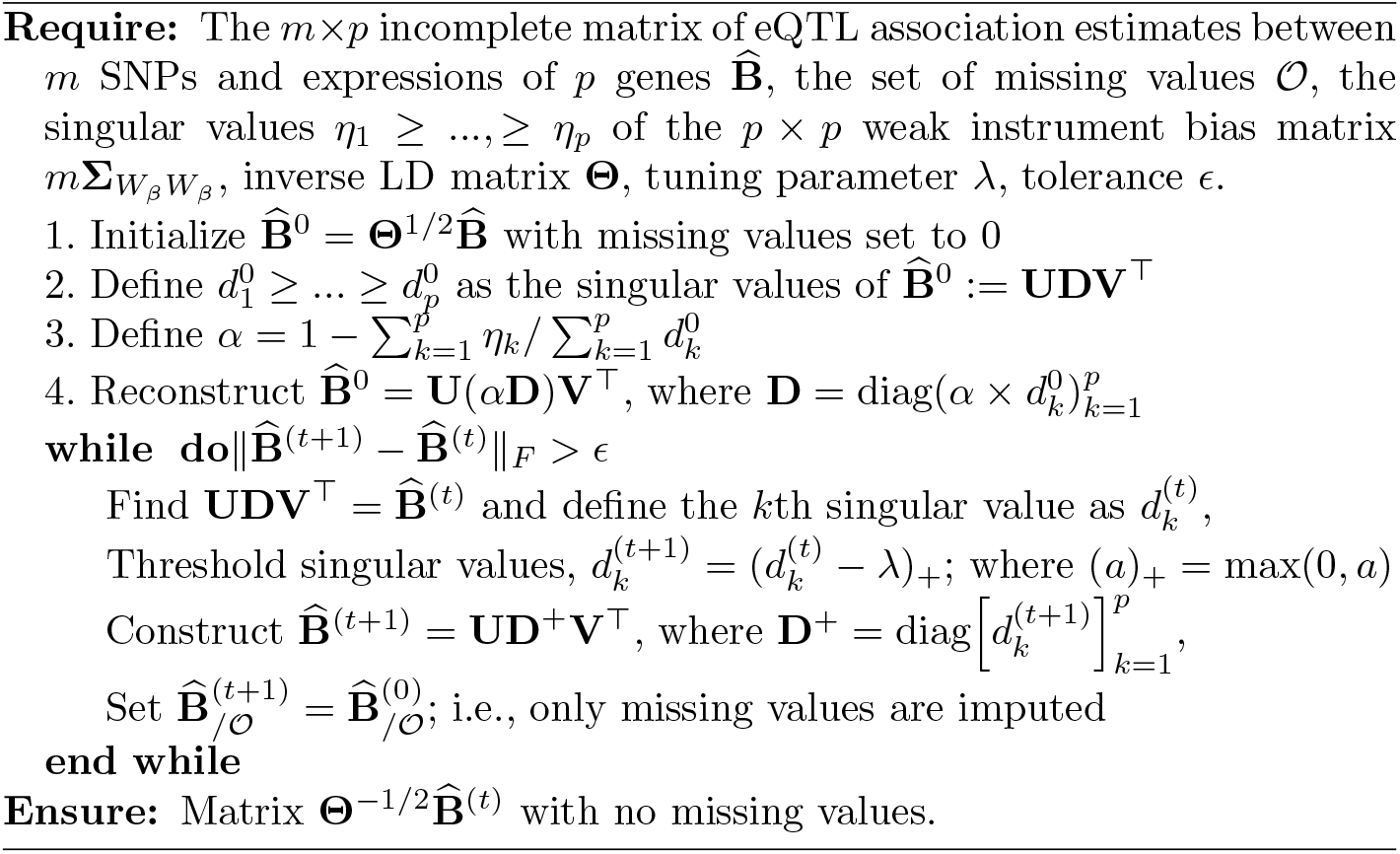

After imputating the missing SNP-expression association estimates, the full set of candidate IVs 𝒮_∪_ is restricted to those that are significant in a joint test of association. Let 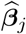 be the *p*-length vector of associations between the *j*th eQTL in 𝒮_∪_ and the expression of *p* genes in a tissue, where 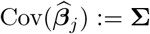 is estimated using the insignificant eQTL effect estimates [34, Method]. The initial candidate set 𝒮_∪_ is restricted to

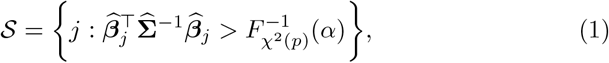

where *α* = 5 × 10^−8^ by default in the HORNET software. The set 𝒮 is further restricted using LD pruning [15, 45] and CHP bias-correction as described in the next section.

### 2.3 Handling linkage disequilibrium

In nearly all applications of MVMR with eQTL data, an estimate of the LD matrix **R** for a set of eQTLs used as IVs is required. There are at least three primary challenges related to the use of eQTLs that are in LD when only individual-level data from a reference panel is available: (i) LD between causal SNPs can induce a correlated horizontal pleiotropy (CHP) bias (see **Supplement Section 2.1**), (ii) imprecise estimates of LD between the eQTLs can lead to underestimated standard errors of the causal effect estimates (**Supplement Sections 2.4** and **2.6**), (iii) direct application of the estimated LD matrix to MR may be impossible because of non positive definiteness and the choice(s) of regularization [3] may not always be clear. An additional challenge which HORNET does not address is the possibility of differences in the LD structure of the population used in GWAS and the LD reference panel. Figure 3 presents results from simulations demonstrating how this can affect inference using MR. In the next three subsections, we describe these challenges in greater detail and present the solutions that HORNET can implement.

#### 2.3.1 Correlated horizontal pleiotropy from LD between eQTLs

CHP can be introduced in eQTL-MVMR if any eQTLs used as IVs in a target locus are in LD with other eQTLs that are not in the IV set. This is a form of confounding that can inflate Type I or II error rates when testing the causal null hypothesis [36, 53]. We account for this CHP by removing IVs in the candidate set 𝒮 that have LD *r*^2^ > *κ* with other SNPs not in this set but within ±2Mb of the boundaries of the locus. A visual example of this process is presented in Panel b of Figure 3. In practice, estimation of LD between eQTLs in the IV set and those outside of it is made using the available LD reference panel. This process will reduce the number of eQTLs available for use in MVMR, since it will remove IVs in LD with neighboring non-IVs, but may provide partial protection against CHP bias.

#### 2.3.2 Inflation from misspecified LD

Mis-specifying the LD matrix corresponding to a set of eQTLs that are used as IVs in eQTL-MR can inflate the statistics used to test the causal null hypothesis [28]. Since individual-level data for the discovery GWAS of the disease phenotype are rarely publicly available, eQTL-MR relies on publicly available reference panels to estimate LD between a set of SNPs using populations which are assumed to be similar to the eQTL GWAS population. This LD matrix can be mis-specified when a reference panel of relatively small size and/or different genetic ancestry is used, making causal inference using standard MR methods such as IVW [4] or principal components adjustment [5] vulnerable to inflated Type I/II error rates [28]. No solution to this problem currently exists for eQTL-MVMR. We demonstrate in this section that this problem is caused by misspecification of the residual degrees of freedom in the standard t-test for statistical inference of a causal effect.

We therefore propose a t-test which is corrected for misspecification of the LD reference panel. Consider a univariable MR model using *m* IVs in which

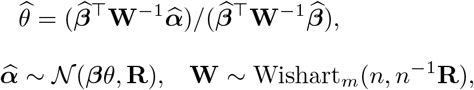

where *n* is the sample size of the LD reference panel. Standard practice to test *H*_0_ : *θ* = 0 compares 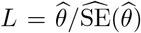 to a t-distribution with *m* − 1 degrees of freedom. This test implicitly assumes that 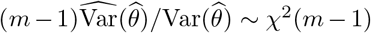, when in fact 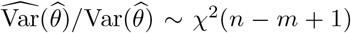 when **W** is treated as random [37]. The statistic *L* does not follow a t-distribution since the residual degrees of freedom is misspecified. However, 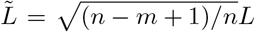 does follow a t-distribution with *m* − 1 degrees of freedom. We therefore use the statistic 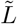 to test *H*_0_ : *θ* = 0 instead of *L*. It follows from the definition of 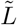 that 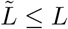, which implies that it may be less powerful than *L*, but should also control the Type I error rate or *L* at the nominal level.

#### 2.3.3 Non-positive definite LD matrix

When using a reference panel to estimate LD between a set of eQTLs that may be used as IVs in eQTL-MVMR, the raw estimate 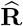 is not guaranteed to be positive definite if the size of the reference panel *n*_ref_ is less than the number of IVs [20]. LD pruning also does not guarantee this issue will always be avoided. In this case, we may not be able to directly use 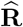 because eQTL-MVMR requires its inverse, which may not exist. Multiple solutions to this problem exist in the literature, with methods either transforming the IV set [5, 38, 57] or directly applying regularization to 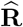 [7]. We allow users to either apply regularization to 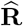 by a scalar factor which achieves positive definiteness with minimal perturbation based on [8], or users may apply LD pruning.

### 2.4 Estimating causal effects

HORNET performs multivariable MR (MVMR) in locus by locus across the genome. Standard causal inference from MVMR is based on the P-value corresponding to the estimated causal effect. We apply this inference and include two additional criteria to prioritize genes based on their significance and estimated causal effect size. These criteria are the (i) locus R-squared, measuring the total contribution of gene expression to phenotypic variation, and (ii) Pratt index [2]. The HORNET software uses MRBEE [34] to estimate causal effects in a set of genes screened as positive by GScreen, which is introduced in the next subsection. MRBEE performs robust multiple regression and so the corresponding variance explained R-squared values can be used to approximately represent the degree of model fit in a locus. We demonstrate in the **Supplement** that the locus R-squared is only equal to the true heritability explained when the power to detect each causal eQTL is 1. The Pratt index is gene-specific in a single locus and is used to represent the gene-specific proportion of variance explained in MVMR. Each locus will have one R-squared value and each gene in the locus will have its own Pratt index value, the sum of which across all genes in the locus is theoretically the locus R-squared value. We introduce the locus R-squared and gene-specific Pratt index values as imperfect measurements of quantities that are generally of interest when applying HORNET, and assert that the MVMR literature currently lacks any measurement which intends to capture what these two do.

#### 2.4.1 Screening genes

We stated in the previous section that each gene in a locus is first screened for evidence of causality then, if passing the screen, their causal effects are estimated using MRBEE. In this section, we briefly introduce the motivation for and execution of the screening process. In a locus of approximately 2Mb, many genes may be present (e.g., upwards of 30). Given the restrictions placed on the structure of cis-eQTL data mentioned in Section 1, the curse of dimensionality may be frequently encountered, making direct estimation of all causal effects in a locus by MRBEE challenging. We therefore propose to first screen all genes in a locus using a variable selection penalty to reduce the dimensionality of MVMR (see [17], [59]). This step will automatically select a relatively small subset of genes with the strongest evidence of direct causality of the outcome. We then apply MRBEE only to the selected genes passing this screening step. We use a new method called GScreen which approximates median regression using the methods of [25] and applies the unbiased SCAD variable selection penalty [17]. Section 4 of the **Supplement** provides more details about the GScreen method and its performance in simulation and application to real data.

### 2.5 Simulations

We performed three separate simulations to assess the performance of missing data imputation, inflation in eQTL-MR, and inflation-correction methods. The setup of each simulation and a discussion of the results they produced are described in the next three subsections.

#### 2.5.1 Imputing missing data

In the missing data simulation, we used summary statistics from eQTL GWAS for 9 genes on chromosome 1 produced from 236 non-Hispanic White individuals. We restricted the eQTLs used to only those within ±2Mb of the transcription start site (TSS) of one of the genes, producing 526 fully observed eQTLs. We then set the Z-statistics for eQTL-gene pairs in which the eQTL was >1Mb from the TSS as missing and evaluated four methods of imputation: (i) MV-Imp, which was the matrix completion approach outlined in Algorithm 1, (ii) imputation of missing values with 0s, (iii) soft impute [35], and (iv) imputation based on the multivariate normal distribution. For each simulation, the true LD correlation matrix **R** between the 526 eQTLs had a first order autoregressive structure with correlation parameter 0.5. The matrix of measurement error correlations 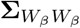 was estimated from all SNPs in the 1Mb window with squared Z-statistics for all eQTL associations less than the 95th quantile of a chi-square distribution with one degree of freedom. This follows the procedures used in practice [34, 61].

In simulation, our multivariate imputation method outlined in Algorithm 1 has smaller estimation error than imputation with all zero values or the traditional soft impute method [35]. Estimation error in this setting is defined as the difference between true and imputed values. Since there is currently no other way to address missing data in eQTL-MVMR, zero-imputation, soft impute, and imputation based on the multivariate normal distribution are three straightforward alternatives to our proposed imputation approach. We demonstrate in Section 1.4 of the **Supplement** and Panel b of Figure 2 that imputing missing data using our algorithm can produce up to 2-4x increases in power vs excluding eQTLs with any missing associations as IVs.

**Fig. 1.**
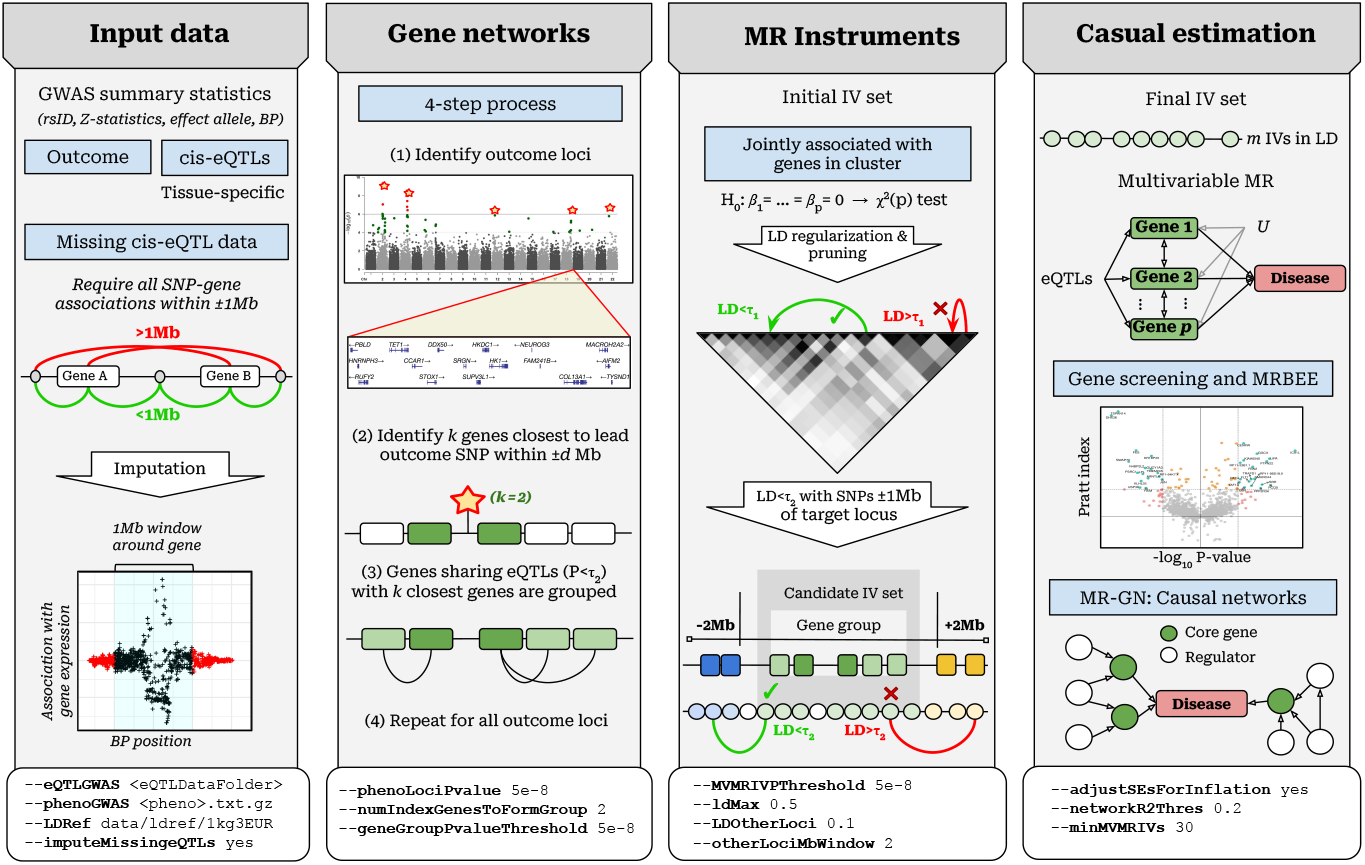
Flowchart illustrating genome-wide causal gene searches using HORNET. Example options given to flags that the command line version of HORNET uses are at the bottom of each panel. In the ‘Input data’ section, ±1Mb is used because it is standard in many publicly available data such as GTEx [10] and eQTLGen [55]. The HORNET software is available from https://github.com/noahlorinczcomi/HORNET

**Fig. 2.**
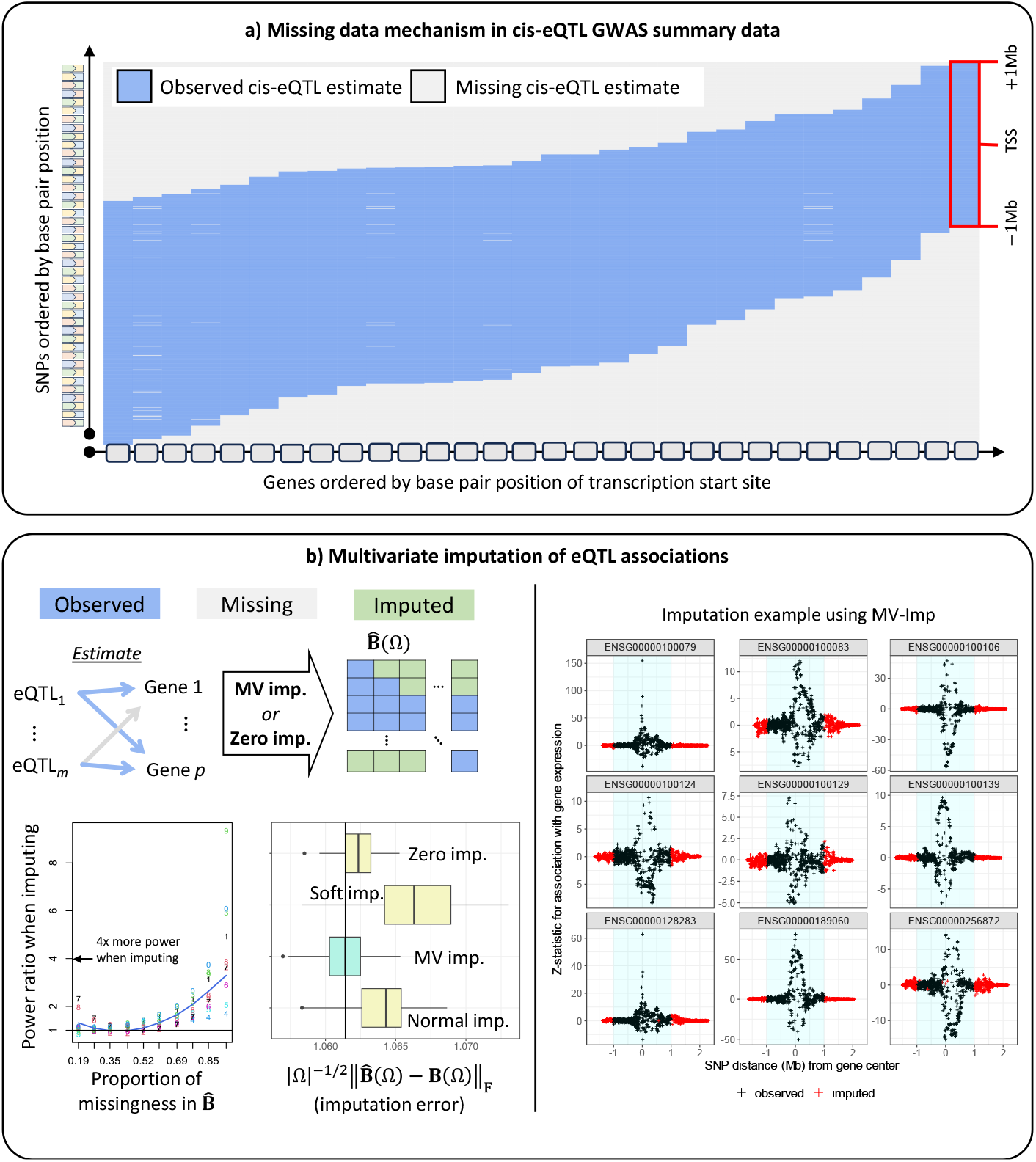
This figure illustrates the mechanism in summary cis-eQTL GWAS data that leads to missing data in eQTL-MVMR and how this missing data can be addressed using imputation. a) Only SNP-gene pairs within a defined distance have association estimates present in cis-eQTL summary data. This figure demonstrates this by displaying the available data for SNPs and genes ordered by their chromosomal position using data from the eQTLGen Consortium [54]. b) (left) Visual display of the pattern of missing in the design matrix 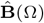 used in eQTL-MVMR. Imputation can be performed by setting missing values to be 0 (‘Zero imp.’) or by applying the low-rank approximation (‘MV imp.’) to 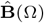 described in Algorithm 1. ‘Soft impute’ is the soft imputation method of [24] and ‘Normal imp.’ is a gene-pairwise imputation method based on the multivariate normal distribution, more fully described in the **Supplement**. |Ω| is the total number of missing values in a simulation performed using real data in the *CCDC163* gene region. These data were GWAS summary statistics of gene expression in blood tissue measured in 236 unrelated non-Hispanic White individuals. Full details of this simulation are presented in the **Supplement**. (right) An example of the MV imp. method applied to summary data for 9 genes on chromosome 22 using cis-eQTL data from the eQTLGen Consortium [54].

#### 2.5.2 Inflation in eQTL-MR

In the simulation to demonstrate inflation in eQTL-MR, the true LD matrix **R** for 500 eQTLs had a first order autoregressive structure with correlation parameter 0.50 and was estimated by sampling from a Wishart distribution with varying degrees of freedom equal to the reference panel sample size. In each simulation, true eQTL and disease standardized effect sizes were drawn from independent multivariate normal distributions with means 0 and covariance matrices **R**. We then applied LD pruning [15, 45] at the threshold *r*^2^ < 0.3^2^ to restrict the IV set used in univariable MR. We performed MR using univariable IVW [4] and the Type I error rate was recorded using both the standard test statistic *L* and the adjusted statistics 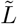 introduced in Section 2.3.2. The Type I error rate was based on tests of the causal null hypothesis. Panel C in Figure 3 demonstrates that LD reference panels that contained genotype information for less than 3,000 individuals inflated the false positive rate in eQTL-MVMR using the standard test statistic *S*. When the reference panel contained 500 individuals, the false positive rate approached 0.25 using *S*. As a comparison, the largest population-stratified sample of individuals in the 1000 Genomes Phase 3 reference sample [9] is 652 and the smallest is 347. Using our adjusted test statistic 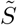, the Type I error rate was controlled at the nominal level for LD reference panels of any size, providing support that this method of hypothesis testing may not have inflated Type I error.

**Fig. 3.**
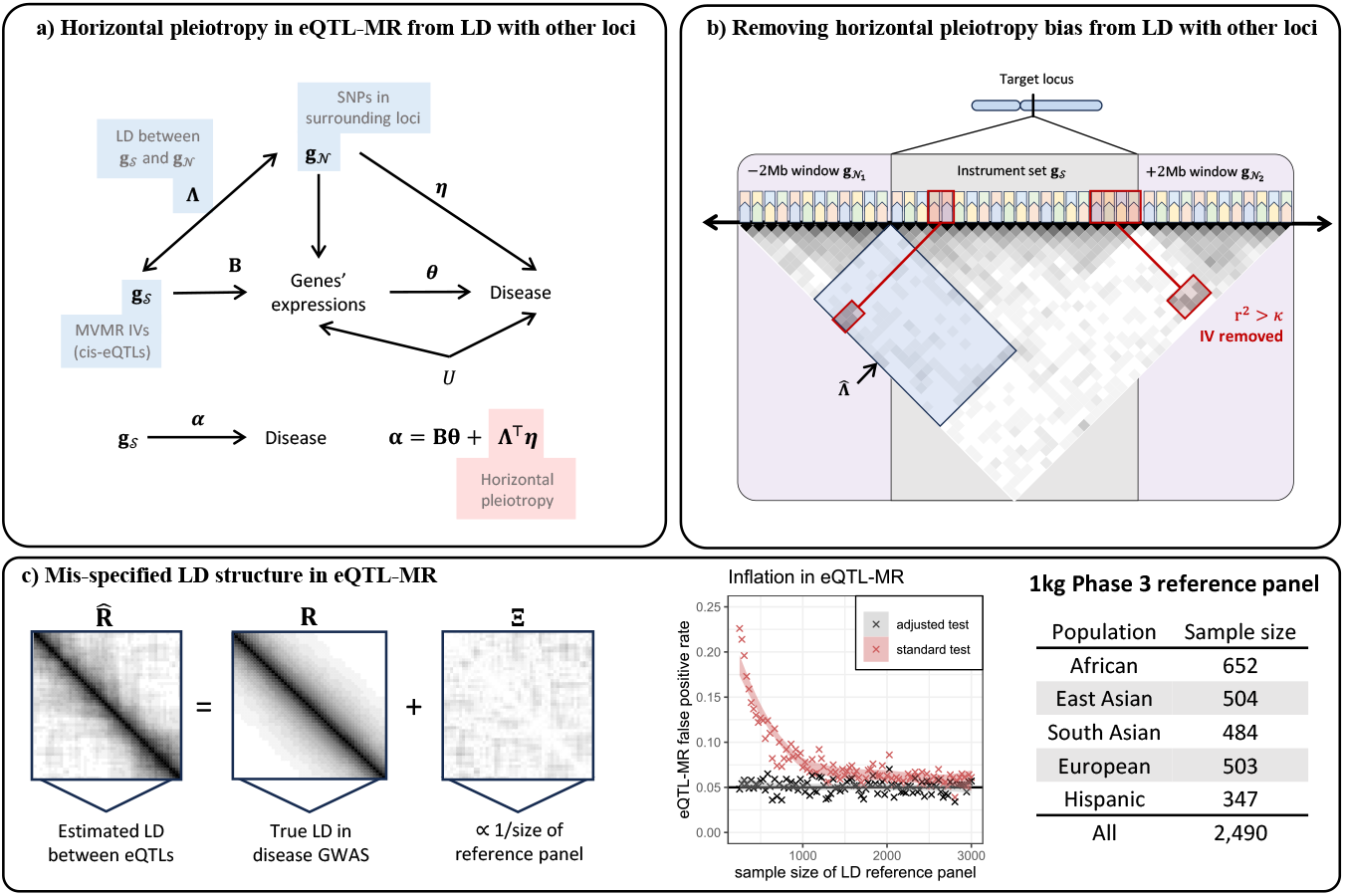
This figure illustrates the adjustments for CHP and inflation that are introduced when the eQTLs used in MR are in LD and researchers only have access to relatively small reference panels. a) The goal of eQTL-MVMR is to estimate ***θ***, which may be subject to bias when **Λ** and ***η*** are each nonzero. b) This is the CHP-adjustment procedure described in Section 2.3.1. c) Results in the panel entitled ‘Inflation in eQTL-MR’ are from simulation in which the true LD matrix had dimension 500 *×* 500 and an AR1 structure with correlation parameter 0.5. We applied LD pruning at the threshold *r*^2^ < 0.3^2^. In this simulation, we repeatedly drew an estimate of the LD matrix from a Wishart distribution with degrees of freedom found on the x-axis. The R code used to perform this simulation is available at https://github.com/noahlorinczcomi/HORNET.

## 3 Implementation

### 3.0.1 Software

HORNET requires GWAS summary statistics for gene expression and a disease phenotype and an LD reference panel. LD estimation from a reference panel for a set of eQTLs is made using the PLINK software [41], which requires the presence of `.bim, .bed`, and `.fam files`. eQTL GWAS data must contain a single file for each chromosome and generally should contain summary statistics for all genotyped SNPs within a cis-region of each available gene. These data are available for blood tissue from the eQTLGen Consortium (n=31k) [54] and the GTEx consortium for 53 other tissues (n<706) [10]. To help researchers identify relevant tissues to select in their analyses, we provide a tissue prioritizing tool based on the heritability of eQTL signals. This tool receives a list of target genes from the researcher and returns a ranked list of tissues in which each target gene has the strongest eQTLs using GTEx v8 summary data [10]. See **Supplement Section 4** for additional details and a demonstration of how to use this tool.

The HORNET software exists as a command line program available for Linux, Windows, and Mac machines. Its tutorial is availabe at https://github.com/noahlorinczcomi/HORNET and is introduced briefly in **Supplement Section 5**. By downloading HORNET, users also receive PLINK v1.9 [41] and LD reference panels for European, African, East and South Asian, Hispanic, and trans-ethnic populations from 1000 Genomes Phase 3 (1kg) [9].

By default, our software uses this reference panel from the entire 1kg sample to estimate LD in the eQTL GWAS population, but users can alternatively specify a specific sub-population in 1kg or even use their own LD reference panels.

### 3.1 Real data analysis with schizophrenia

We applied the HORNET methods and software to the analysis of genes whose expression in basal ganglia, cerebellum, cortex, hippocampus, amygdala, and blood tissues cause schizophrenia risk. Schizophrenia GWAS data were from [50], which included 130k European individuals and were primarily from the Psychiatric Genomics Consortium (PGC) core data set. eQTL GWAS data in brain tissue were from [13], which contained GWAS data from European samples of sizes 208 for basal ganglia, 492 for cerebellum, 2,683 for cortex, 168 for hippocampus, and 86 for amygdala tissue. eQTL GWAS data in blood were from the eQTLGen Consortium [54] for 31k predominantly European individuals. We performed analyses with HORNET in all schizophrenia loci with at least one P-value less than 0.005, grouped genes sharing eQTLs with P-values less than 0.001, applied LD pruning at the threshold *r*^2^ < 0.7^2^, and removed SNPs in LD with any IVs in the target locus beyond *r*^2^ > 0.5^2^ in a 1Mb window. Finally, all IVs had a P-value for joint association with gene expression across all tissues which was less than 5×10^−3^ in the test of Equation 1. We performed HORNET in each tissue separately and present the results in Figure 4.

**Fig. 4.**
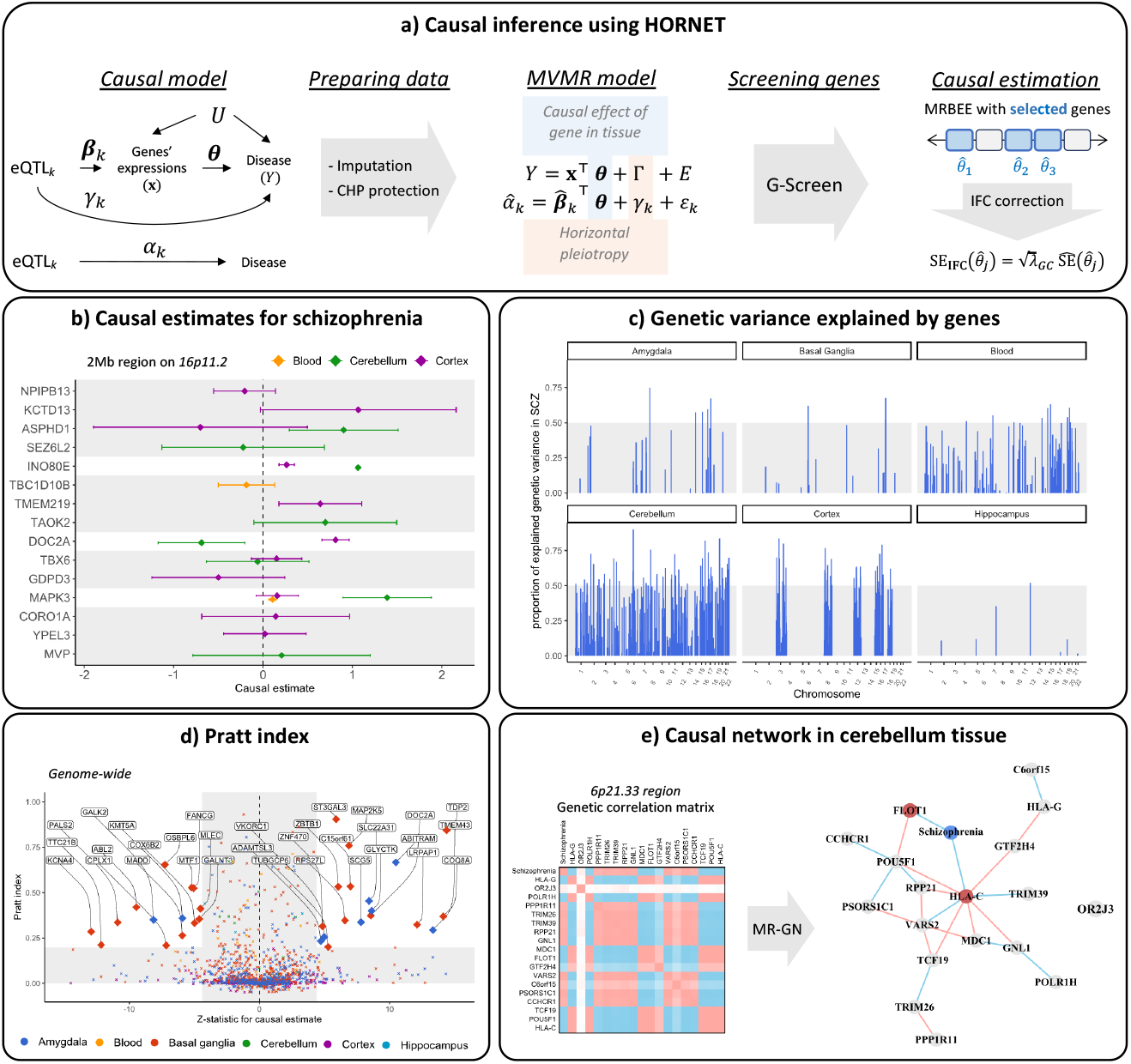
This figure presents the results of using HORNET to search for genes modifying schizophrenia risk when expressed in different tissues. a) Description of the causal model, MVMR model, and estimator. b) Causal estimates for multiple genes in blood, cerebellum, and cortex tissues in the schizophrenia-associated *KCTD13* locus. c) R-squared values from MVMR models fitted across the genome. Areas in which no R-squared values exist either had no genes prioritized by GScreen or had insufficient eQTL signals to perform MVMR. d) Pratt index values for all causal estimates made for all tissues. Pratt index values outside the range of (−0.1,1) are not shown. This may happen because of large variability in univariable MR estimates for some loci. e) Estimated gene regulatory and schizophrenia causal network for 18 genes in the schizophrenia-associated *FLOT1* locus of the HLA complex graphical lasso [18].

Figure 4 uses the data described above to provide examples of the primary results produced by genome-wide analysis with HORNET, including causal estimates for prioritized genes, genome-wide R-squared and Pratt index values for each tissue, and an estimated sparse regulatory network of genetic correlations using graphical lasso [18]. These results show that locus R-squared values can exceed 0.50 for many loci, suggesting that SNP associations with schizophrenia in these loci may be primarily explained by gene expression in brain tissue (Panel c). For example, 17.2% of genetic variation in schizophrenia in the *KCTD13* locus is explained by the expression of genes in blood tissue, 75.2% in the cerebellum, and 59.4% in the cortex. In this locus, we observed that expression of the *INO80E* gene in the cortex increased schizophrenia risk (*P* = 2.1 × 10^−9^), but that the specific schizophrenia variation attributable to this effect was small (Pratt index=0.09). Alternatively, expression of the *DOC2A* gene in the cortex was strongly associated with increased schizophrenia risk (*P <* 10^−50^) and also had a relatively large Pratt index value of 0.67 (Panels b and d), suggesting that *DOC2A* is potentially a better gene target than *INO80E* in the cortex.

We attempted to better understand the complex regulatory network that exists in the human leukocyte antigen (HLA) complex of 6p21.33 [30]. Genetic variants in this region are highly associated with risk of schizophrenia [11, 23, 27, 27] and many other traits such as brain morphology [6], autism spectrum disorder [1], and Type II diabetes [56]. The HORNET software applied graphical lasso [18] to the matrix of imputed marginal Z-statistics to uncover regulatory relationships between 18 genes in this locus and their pathways of causal effect on schizophrenia risk when expressed in cerebellum tissue. These results suggest a densely connected gene regulatory network in which the *HLA-C* gene is a so-called ‘regulatory hub’ [14, 58]. The *HLA-C* gene is directly associated with the regulation of 8 other genes and is indirectly associated with the regulation of all genes in the locus except *OR2J3*. Only *HLA-C* and *FLOT1* have direct causal effects on schizophrenia risk, and all other 15 peripheral genes (*OR2J3* excluded) have causal effects on schizophrenia that only are mediated by *FLOT1* and/or *HLA-C* expression.

## 4 Discussion

Existing methods for finding causal genes using multivariable Mendelian Randomization (MR) with GWAS summary statistics are generally vulnerable to bias and inflation from missing data, misspecified LD structure, and confounding by other genes. Equally, no flexible and comprehensive set of computational tools to robustly perform this task current exists. We introduced a suite of statistical and computational tools in the HORNET software that addresses these common challenges in multivariable MR using eQTL GWAS data. HORNET can generally provide unbiased causal estimation and robust inference across a range of real-world conditions in which existing methods in alternative software packages may not. HORNET is a command line tool that can be downloaded from https://github.com/noahlorinczcomi/HORNET, where users will also find detailed tutorials demonstrating how to use HORNET.

## Data Availability

All data generated by the study is available at our Github page.

https://github.com/noahlorinczcomi/HORNET

https://github.com/noahlorinczcomi/HORNET_AD

## 5 Acknowledgements

This work was supported by [grant numbers HG011052, HG011052-03S1] (to X.Z.) from the National Human Genome Research Institute (NHGRI). NLC was partially supported by [grant number T32 HL007567] from the National Heart, Lung, and Blood Institute (NHLBI).

