## Supplement for "HORNET: Tools to find genes with causal evidence and their regulatory networks using eQTLs"

**3 Estimating bias-correction terms** **31**

**4 Gene Selection** **32**

**5 Prioritizing tissues** **33**

**6 Software** **36**

### 1 Missing data

#### 1.1 Demonstration of missingness

As mentioned in the main text, the set of instrumental variables (IVs) used in multiple exposure Mendelian Randomization (MVMR) is the union set of exposure-specific IV sets. In summary data from cis-eQTL GWAS in which each exposure is the expression of gene, not all SNPs are tested for an association with each gene. Generally, only SNPs within ±1Mb of a gene are tested for an association with the expression of that gene. This means that the union set may contain at least some SNPs for which there is no estimate of association between them and each gene in a locus under study. Visual representations of this are displayed in Figures 1 and 2. To avoid introducing missing data by using the union set of gene-specific IV sets in MVMR, one may consider using the intersection set of gene-specific IV sets, guaranteeing no missing data. However, for a locus containing a moderately large number of genes (e.g., ∼10 or more), the intersection set may actually be of very small size or even empty. This could respectively introduce a $p > n$ scenario or even prevent MVMR from performed.

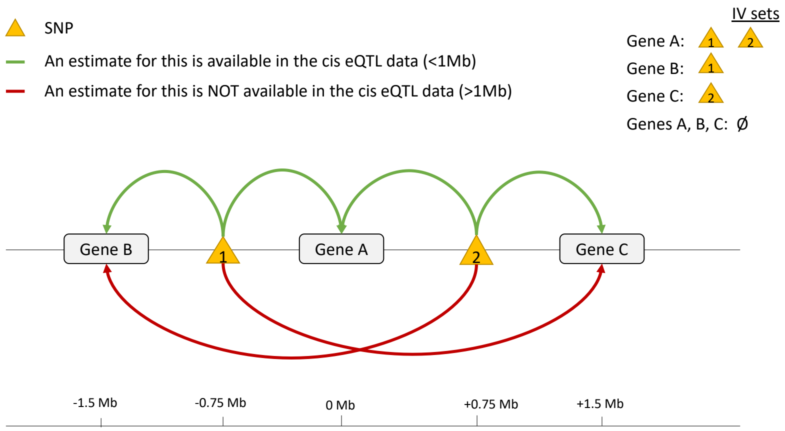

**Fig. 1** This is an example representation of the data that is available in the summary-level eQTLGen [33] and GTEx [9] cis-eQTL public data sets. Only associations between SNPs and the expression of genes within ±1Mb of those SNPs have estimates included in the available data. Since in multivariable MR, we select as the IV set the union of gene-specific IV sets, this union set may contain no SNPS with association estimates observed for all genes in a group. Put another way, the intersection of all gene-specific IV sets that is restricted only to SNPs with no non-missing values may be empty.

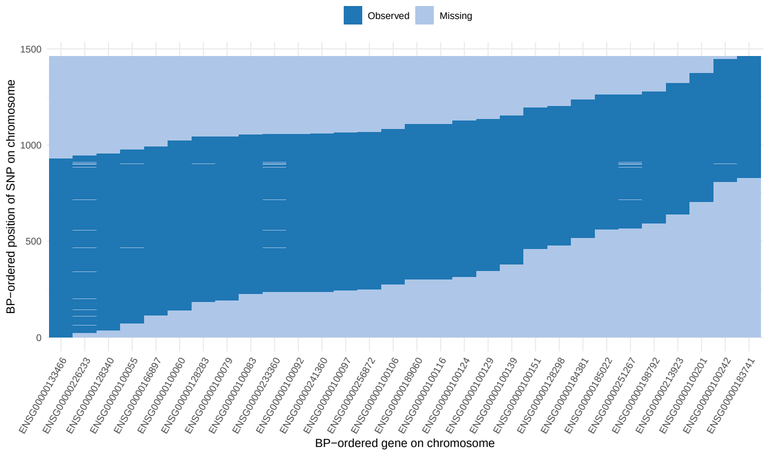

**Fig. 2** Determinations of missing values for each SNP (in order by BP position on y-axis) that was used in multivariable MR with the 30 genes ordered by BP position on CHR 22 (x-axis), as an example. These genes were grouped using the procedure described in **Methods**. It was stated in **Methods** that the nature of the *cis*-eQTL data is such that only SNPs within +-1Mb of a gene center have estimates of association with the gene expression available in the data. In our analyses, we included multiple genes in causal estimation. Denote the set of SNPs used as IVs in multivariable MR for a group of genes of size $p_k$ as $\mathcal{M}_k$, which is the union set $\cup_{\ell=1}^{p_k}\mathcal{M}_k^{\ell}$ of the gene-specific IV sets $\mathcal{M}_k^1 ... \mathcal{M}_k^{p_k}$. This union set is the set of SNPs whose ordered positions are on the y-axis. As Figure 1 demonstrated, restricting this union set to only non-missing association estimates between each SNP and gene expressions may make the set empty. In the figure above, this scenario would correspond to being unable to draw any horizontal line through the plot such that the line never touches a 'Missing' area.

| Tissue | Minimum % missing eQTL associations across genes in a locus | | Maximum % missing eQTL associations across genes in a locus | | Locus size (Mb) | | |
|---|---|---|---|---|---|---|---|
| | Minimum across all loci | Mean across all loci | Mean across all loci | Maximum across all loci | Minimum | Maximum | Mean |
| Basal ganglia | 0.00 | 0.11 | 0.16 | 0.60 | 1.04 | 3.32 | 1.81 |
| Blood | 0.00 | 0.20 | 0.31 | 0.72 | 0.26 | 3.89 | 1.95 |
| Cerebellum | 0.00 | 0.06 | 0.10 | 0.58 | 0.35 | 3.25 | 1.58 |
| Coronary artery | 0.00 | 0.09 | 0.14 | 0.58 | 0.64 | 3.05 | 1.82 |
| Cortex | 0.00 | 0.05 | 0.08 | 0.59 | 0.33 | 3.87 | 1.57 |
| Hippocampus | 0.00 | 0.12 | 0.17 | 0.45 | 0.82 | 2.91 | 1.71 |
| Lung | 0.00 | 0.08 | 0.12 | 0.55 | 0.51 | 3.27 | 1.59 |
| Spinal cord | 0.00 | 0.06 | 0.09 | 0.49 | 1.23 | 2.61 | 1.80 |

**Fig. 3** This figure presents a high-level summary of the rates of missing eQTL associations in gene groups formed while applying HORNET to the study of schizophrenia (see main text). Values in the first four columns after tissue type correspond to missingness rates; values in the final three columns correspond to the sizes, from the smallest base pair position of an eQTL used as an IV, to the largest, of loci analyzed by HORNET. Missingness rates are first aggregated from the gene level to the locus level, then again from the locus level to the genome level. For example, the '0.00' value in the first row and second column indicates that the smallest rate of missingness observed for any gene that was analyzed by HORNET in basal ganglia tissue was 0.00, the next column indicates the mean rate of missingness across all loci analyzed by HORNET in the same tissue, and so on. eQTL GWAS data for basal ganglia, cerebellum, cortex, hippocampus, and spinal cord tissues were from [10]; coronary artery and lung tissue data were from [8]; blood tissue data were from [33]. The complete set of commands given to HORNET to perform these analyses is available at https://github.com/noahlorinczcomi/HORNET/real_data.

#### 1.2 Support from cis-eQTLs in a larger window

Since most publicly available summary data from cis-eQTL GWAS contain association estimates between SNPs and the expression of genes within ±1Mb of each them, we wanted to better understand the pattern of association between gene expression and SNPs >1Mb away. For this, we used individual-level data from 236 non-Hispanic unrelated White individuals. We estimated associations between gene expression and all available SNPs within ±5Mb using the TensorQTL pipeline [32]. Displayed in Figure 4 are these association estimates for 25 genes on chromsome 1 that had eQTLs with corresponding P-values less than $5 \times 10^{-8}$. These results demonstrate that, on average, the most significant eQTL signals are near the transcription start site and that significant eQTLs are unlikely to be observed outside of a 1Mb window but within 5Mb.

#### 1.3 Multivariate Imputation

##### 1.3.1 Procedure

In this subsection, we describe the procedure that we used to impute missing data in the set $\cup_{\ell=1}^{p}\mathcal{M}^{\ell}$ that is the union of $p$ gene-specific IV sets each denoted as $\mathcal{M}^{\ell}$. Our imputation method is similar to the soft imputation method using matrix completion [22] but corrects for measurement error in the eQTL GWAS

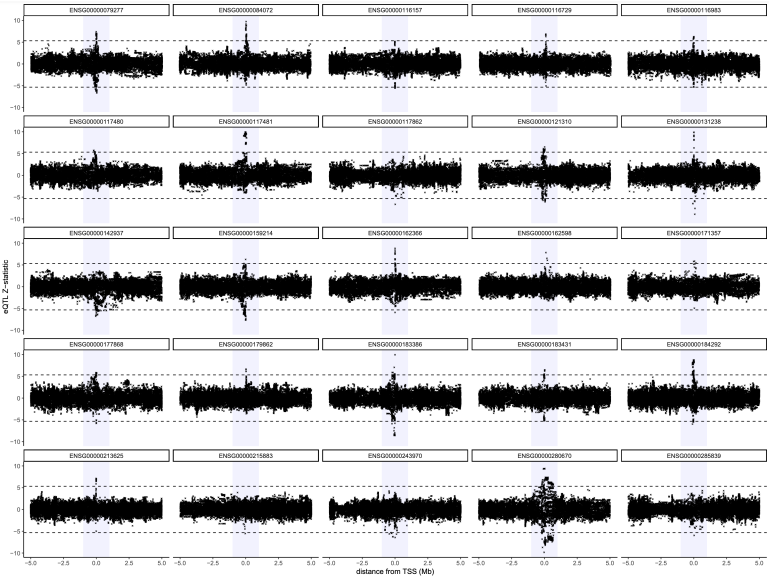

**Fig. 4** Displayed are Z-statistics for eQTLs that are within ±5Mb around 25 genes on chromosome 1 in blood tissue and non-Hispanic unrelated White individuals. The shaded blue regions represent the ±1Mb region around the transcription start site (TSS) for each gene. Horizontal dashed lines represent $\pm F^{-1}(1 - 5 \times 10^{-8})$, where $F^{-1}(\alpha)$ is the quantile function of the standard normal distribution evaluate at $\alpha$. These results indicate that eQTL Z-statistics are highly likely to be considered not genome-wide significant, i.e., to have a corresponding P-value greater than $5 \times 10^{-8}$ outside of the ±1Mb window from the TSS.

and accounts for LD structure. 'Measurement error in the eQTL GWAS' here refers to the nonzero variance of $\hat{\beta}_{j\ell}$, the *estimated* association of the $j$th SNP with the $\ell$th gene in a select tissue. Only when $\hat{\beta}_{j\ell} = \beta_{j\ell}$, the *true* association, is there no measurement error in $\hat{\beta}_{j\ell}$. Let $\mathbf{\Sigma}_{W_\beta W_\beta}$ denote the $p \times p$ variance-covariance matrix of $\hat{\boldsymbol{\beta}}_j$, the $p$-length vector of associations between the $j$th SNP and $p$ genes in a locus. Let $\widehat{\mathbf{B}}$ be the $m \times p$ matrix of estimated associations between $m$ SNPs and the expression of $p$ genes, $\mathbf{B}$ denote the corresponding matrix of true associations, and $\mathcal{O}$ be the set of non-missing values in $\widehat{\mathbf{B}}$, of which there are $|\mathcal{O}|$.

The main principle of soft imputation is to iteratively apply soft thresholding to the singular values of $\widehat{\mathbf{B}}$ until convergence. Since $\widehat{\mathbf{B}}$ contains missing values, we first impute the missing values in $\widehat{\mathbf{B}}$ with 0, a reasonable estimate of their true value given the results from individual-level data presented in Figure 4. Our matrix completion algorithm is presented in Algorithm 1 in the main text. This algorithm modifies the traditional soft impute method [22] by subtracting the singular values of $\mathbf{\Sigma}_{W_\beta W_\beta}$ from the singular values of an initialized $\widehat{\mathbf{B}}$. This method requires the tuning parameter $\lambda$ to be used in soft thresholding and will only accept solutions in which the rank of the imputed matrix is less than a user-defined value. Below, we evaluate the performance of this imputation method in simulation in Figure 6 and provide some examples using real data in Figure 7.

##### 1.3.2 Simulations with generated data

First, we simulated true associations between $m = 150$ SNPs and $p = 10$ genes, which formed the matrix $\mathbf{B}$. Next, we randomly drew association estimates $\hat{\mathbf{B}}$ from the matrix normal distribution $\mathcal{N}(\mathbf{B}, \mathbf{R}, \mathbf{\Sigma}_{W_\beta W_\beta})$, where $\mathbf{R}$ is the matrix of LD correlations between the 100 SNPs. In our simulations $\mathbf{R}$ had a first-order autoregressive structure with correlation parameter $\rho$ which was in the set $\{0.0, 0.1, ..., 0.8, 0.9\}$. The matrix $\mathbf{\Sigma}_{W_\beta W_\beta}$ representing measurement error covariance between the rows of $\hat{\mathbf{B}}$ had a Toeplitz structure and was multiplied by the factor $\sqrt{m} \log p \approx 5.3$. We then applied our matrix completion algorithm to these data, searching over a grid of $\lambda$ parameter values and fixing the maximum acceptable rank of the solution at 5.

The simulation results in Figure 6 demonstrate that our imputation method well-approximates the true underlying distribution of the observed association values when the true mean of the missing association values is 0, and that LD structure does not appear to affect these results. An example of the imputation for a single case is presented in Figure 7. Results from individual level data presented in Figure 4 demonstrate that the true mean is likely to be 0 for almost all areas outside of the $\pm$1Mb window of a gene's center. Results from real data presented in Figure 8 demonstrate that this imputation method can capture the variance in association estimates at the boundaries of the observed windows well, and that association estimates further from the gene center approach 0 with decreasing variance. The data used in these results are described in the caption of Figure 8.

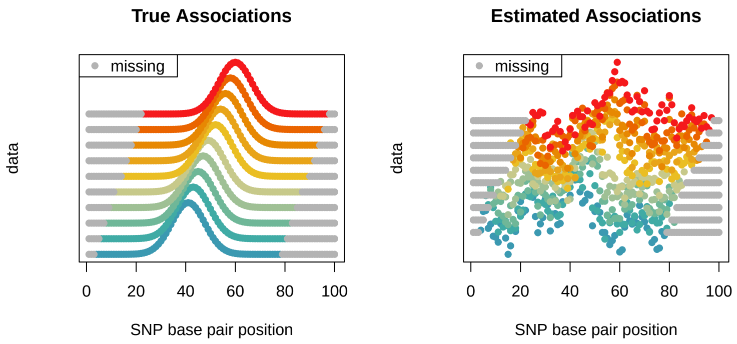

**Fig. 5** This figure displays the data we generated to perform simulation using our eQTL imputation method described in Algorithm 1 in the main text. Positions on the x-axis correspond to unique SNPs. Each gene is represented by a different color, where gray always represents missing values. For each gene, y-axis values are arbitrary but the relative magnitude corresponds to the magnitude of association with the SNP at that base pair position with the expression of the specific gene. The base pair locations of missingness for each gene depend on the base pair position of the gene center, which is located at the peak of its distribution. Gene centers/distribution peaks are staggered for each gene to replicate the real data. The left panel displays true association values and the right panel displays an example of estimated association values.

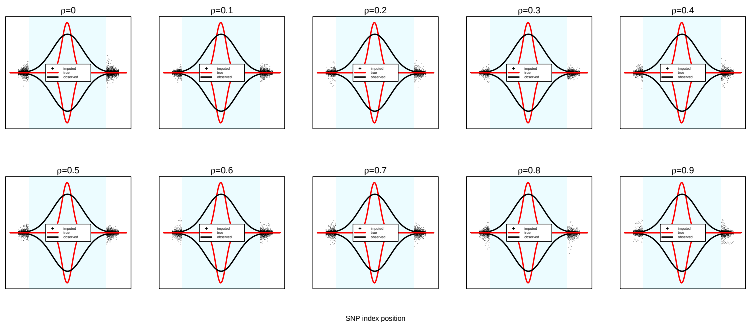

**Fig. 6** These are the results of simulations in which LD structure among the 150 SNPs varied from an AR1 structure with correlation paramater $\rho = 0$ to $\rho = 0.9$. Y-axis display the relative strength of association between a SNP indexed on the x-axis and the expression of the first gene of 10 in simulation. The true distributions of the true and observed associations are respectively represented by red and black lines. All imputed values across all 100 simulations are represented by black points.

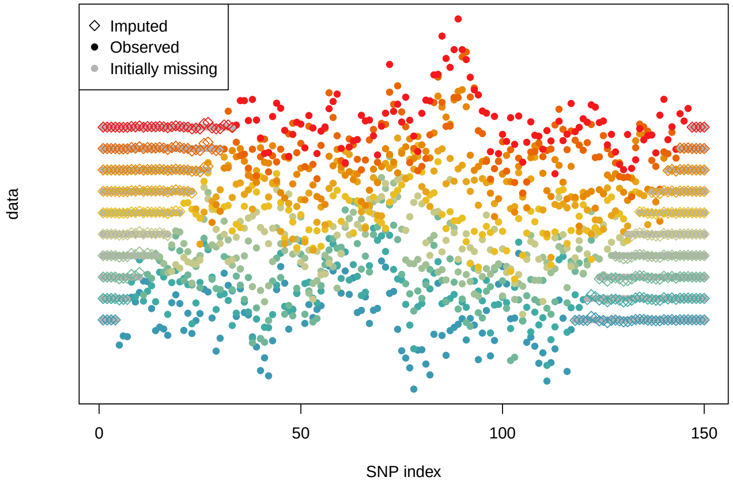

**Fig. 7** These are the results of simulations which are described in the text above this figure when $\mathbf{R}$ has an AR1 structure with correlation parameter $\rho = 0.5$. These results demonstrate that our imputation method well approximates the true underlying association values when the data is missing, which in this simulation are each 0. Observed values are equal to the true values plus measurement error from finite GWAS sample size.

##### 1.3.3 Simulations with real data

We also performed simulations using GWAS summary statistics from 236 unrelated non-Hispanic White individuals whose gene expression in blood tissue were measured. These data provided estimates of association between SNPs and the expression of genes whose transcription start sites (TSSs) were within ±5Mb of them. We selected the *CCDC163* locus in the 1p34.1 region and considered the eight genes closest to *CCDC163* to form the completely observed matrix of Z-statistics $\mathbf{Z}$ which was of dimension $526 \times 9$. For each gene, we then set all GWAS association estimates outside of a ±1Mb region of the TSS to be missing. We set the true LD correlation matrix ($\mathbf{R}$) between the 526 SNPs to be of a first-order autoregressive structure with correlation parameter 0.50, and true genetic covariance ($\mathbf{S}$) between genes to that which was observed empirically from $\mathbf{Z}$. For each of 50 iterations, we performed the following steps:

1. Draw $\mathbf{E}^*$ from Normal$(\mathbf{0}, \mathbf{S}, \mathbf{R})$
2. Calculate $\mathbf{Z}^* = \mathbf{Z} + \mathbf{E}^*$
3. Set values in $\mathbf{Z}^*$ from the set $\Omega$ to be missing
4. Impute missing values to form complete matrix $\mathbf{Z}_I^*$
5. Calculate $\|\mathbf{Z}_I^* - \mathbf{Z}\|_F$.

The results of this procedure are presented in Panel C of Figure 2 in the main text and show comparisons between four imputation methods: (i) the matrix completion method we introduced in Algorithm 1 in the main text ('MV imp.'), (ii) imputation of missing values with 0's ('Zero imp'), (iii) imputation using the soft impute method ('Soft impute') [16], and (iv) an imputation method based on the multivariate normal distribution ('Normal imp.'. We now briefly describe the Normal imp. method, since the Zero imp. is obvious, MV imp. is described above, and soft impute is described in [16]. Consider the following models for two SNPs for which $n$ individual genotypes have been sampled and placed into the $n \times 1$ vectors $\mathbf{g}_1$ and $\mathbf{g}_2$, for which $\mathbf{g}_1^\top \mathbf{g}_1 = n$ and $\mathbf{g}_2^\top \mathbf{g}_2 = n$. Let $\mathbf{x}$ be the corresponding $n$-length vector of gene expression measurements standardized to have variance 1. The Z-statistics for association between the two SNPs and gene expression are $Z_1 = \mathbf{g}_1^\top \mathbf{x}/\sqrt{n}$ and $Z_2 = \mathbf{g}_2^\top \mathbf{x}/\sqrt{n}$, and it immediately follows that $\mathrm{E}(Z_2|Z_1) = r_{12}Z_1$ where $r_{12}$ is the LD correlation between the two SNPs. This can be shown to easily extend to a multivariate case in which $\mathrm{E}(\mathbf{z}_2|\mathbf{z}_1) = \mathbf{R}_{12}^\top \mathbf{R}_{11}^{-1}\mathbf{z}_1$ where $\mathbf{z}_2$ is a vector of Z-statistics of arbitrary length, $\mathbf{z}_1$ is correspondingly similar for a different set of SNPs, $\mathbf{R}_{12}$ is the matrix of LD correlations between the two sets of SNPs, and $\mathbf{R}_{11}$ is the matrix of LD correlations between SNPs corresponding to $\mathbf{z}_1$. In practice, where $\mathbf{Z}$ represents a matrix of arbitrary dimensions with missing values, the Normal imp. procedure imputes missing values using their conditional expectation from the most fully observed column vector of $\mathbf{Z}$.

#### 1.4 Power after imputing missing values

As mentioned above and in the main text, current eQTL-MVMR approaches are restricted to using IVs for which associations between all SNPs and target genes have been observed in the summary eQTL GWAS data. In this section, we present the results of a simulation in which we compare the power of our multivariate imputation method and current methods that use only completely observed data for testing the causal null hypothesis. We simulated summary-level data using the same procedure described in Section 1.3.2 but varying proportions of total missingness in the true $100 \times 10$ design matrix $\mathbf{B}$, ranging from 19% to 85%. This was accomplished by varing the proportion of missingness that was present for each gene. We compared the power of the IVW method [5] with correlated IVs when we excluded IVs with any missing to power when we imputed missing using our IFC approach. The full R code used to perform these simulations and generate its results are present at https://github.com/noahlorinczcomi/HORNET/simulations.

These results indicate that using imputed vs fully observed data can result in tests of the causal null hypothesis that are up to approximately 4 times as powerful when the proportion of missingness is large. When the proportion of missingness is moderate around 52%, which is consistent with the results observed in real data analyses (see Figure 3 in Section 1.1), still applying imputation to the observed eQTL summary statistics can result in approximately 1.27x more power. The gains in power continue to become larger after approximately 35% or more of the eQTL associations are imputed.

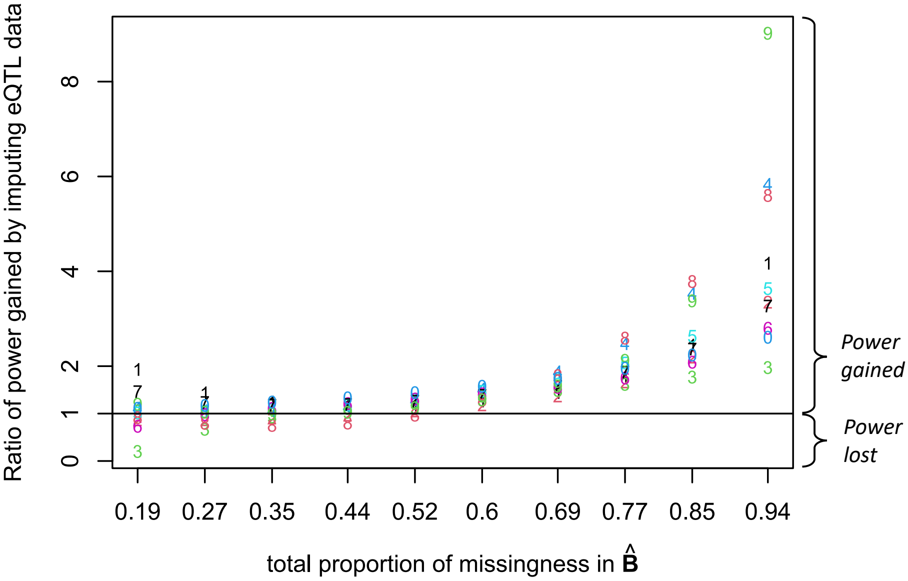

**Fig. 8** This figure displays differences in the power of eQTL-MVMR using IVW with completely observed vs imputed eQTL summary statistics. The x-axis is the proportion of missingness in the working design matrix $\hat{\mathbf{B}}$ and the y-axis the the power when using imputed data divided by the power when using the fully observed data (a 'complete SNP' analysis). 'Power' in this setting refers to the power of rejecting the causal null hypothesis. Each point is a number which indicates the gene. For example, '1' corresponds to the first gene and '0' to the 10th. The horizontal line is placed at 1, below which the complete SNP analysis is more powerful than the analysis using imputed data and above which the converse is true by the factor displayed on the y-axis. Displayed are the power ratios after smoothing power estimates within each analysis type using quadratic regression.

### 2 Accounting for LD in eQTL-MVMR

#### 2.1 CHP bias from LD

##### 2.1.1 Notation

A $m \times p$ matrix $\mathbf{X}$ with normally-distributed elements will be denoted as $\mathbf{X} \sim \mathcal{N}(\boldsymbol{\mu}, \boldsymbol{\Sigma}, \mathbf{R})$, where $\boldsymbol{\Sigma} : p \times p$ represents covariance between columns of $\mathbf{X}$ and $\mathbf{R} : m \times m$ covariance between rows. $\mathbf{X}$ can also be written in vectorized form as $\text{vec}(\mathbf{X}) \sim \mathcal{N}(\text{vec}(\boldsymbol{\mu}), \boldsymbol{\Sigma} \otimes \mathbf{R})$.

Consider two loci (denoted as locus 1 and 2), where locus 1 contains $p_1$ genes that use $m_1$ SNPs as instruments (IVs) and locus two contains $m_2$ SNPs, where all of the $m_1$ SNPs are *cis*-eQTLS for at least one gene in their locus. Denote $\widehat{\mathbf{B}}_1 = (\beta_{ij})_{i,j=1}^{m_1,p_1} : m_1 \times p_1$ and $\widehat{\mathbf{B}}_2 : m_2 \times p_2$ as the GWAS estimates for association with the expressions of the $p_1$ genes in locus 1 and the $p_2$ genes in locus 2. Denote $\widehat{\boldsymbol{\alpha}}_1$ as outcome GWAS estimates for $m_1$ SNPs in locus 1. Assume all GWAS estimates are standardized to have variance 1 and let $\mathbf{R}_1 : m_1 \times m_1$ and $\mathbf{R}_2 : m_2 \times m_2$ denote the true LD correlation matrices for SNPs in loci 1 and 2, where $\mathbf{R}_{12} : m_2 \times m_1$ is the LD correlation matrix between the $m_1$ and $m_2$ SNPs. Define $\mathbf{x}_1 : p_1 \times 1$ as the vector of total gene expression in a tissue for $p_1$ genes in locus 1 and $\mathbf{g}_1 : m_1 \times 1$ as the genotype vector for the $m_1$ SNPs in locus 1.

##### 2.1.2 Models

Consider that the causal effects $\boldsymbol{\theta}$ of the $p_1$ gene expressions in locus 1 on the outcome trait $y$ are of interest and we want to use MR to estimate them. First, we can specify a model for the relationship between $\mathbf{g}_1$ and $\mathbf{g}_2$. Assume that the elements of $\mathbf{g}_1$ and $\mathbf{g}_2$ are approximately normally distributed, or there is some underlying normal distribution from which their realizations are drawn. It follows that $\mathbf{g}_2 = \boldsymbol{\lambda}_{12}^\top \mathbf{g}_1 + \boldsymbol{\epsilon}_2$ where $\boldsymbol{\lambda}_{12} \approx \mathbf{R}_{12}\mathbf{R}_1^{-1}$. Now we specify the following models for the expression values for the $p_1$ genes in locus 1 and their causal effects on the outcome trait:

$$
\begin{aligned}
\mathbf{x}_1 &= \boldsymbol{\gamma}_1^\top \mathbf{g}_1 + \boldsymbol{\gamma}_2^\top \mathbf{g}_2 + \boldsymbol{\epsilon}_1 && (1)\\
&= (\boldsymbol{\gamma}_1^\top + \boldsymbol{\gamma}_2^\top \boldsymbol{\lambda}_{12}^\top)\mathbf{g}_1 + \tilde{\boldsymbol{\epsilon}}_1, && (2)\\
&= \mathbf{B}_1^\top \mathbf{g}_1 + \tilde{\boldsymbol{\epsilon}}_1 && (3)\\
y &= \boldsymbol{\theta}^\top \mathbf{x}_1 + \boldsymbol{\pi}^\top \mathbf{g}_2 + \boldsymbol{\epsilon}_y && (4)\\
&= \boldsymbol{\theta}^\top \mathbf{B}_1^\top \mathbf{g}_1 + \boldsymbol{\pi}^\top \boldsymbol{\lambda}_{12}^\top \mathbf{g}_1 + \tilde{\boldsymbol{\epsilon}}_y && (5)\\
&= \boldsymbol{\alpha}_1^\top \mathbf{g}_1 + \tilde{\boldsymbol{\epsilon}}_y, && (6)
\end{aligned}
$$

where $\tilde{\boldsymbol{\epsilon}}_y$ represents uncorrelated error in a simplified notation. The above results imply that

$$\boldsymbol{\alpha}_1 = \mathbf{B}_1\boldsymbol{\theta} + \boldsymbol{\lambda}_{12}\boldsymbol{\pi}, \qquad (7)$$

where we want to use MR to estimate $\boldsymbol{\theta}$. Figure 9 shows these models in a directed acyclic graph (DAG).

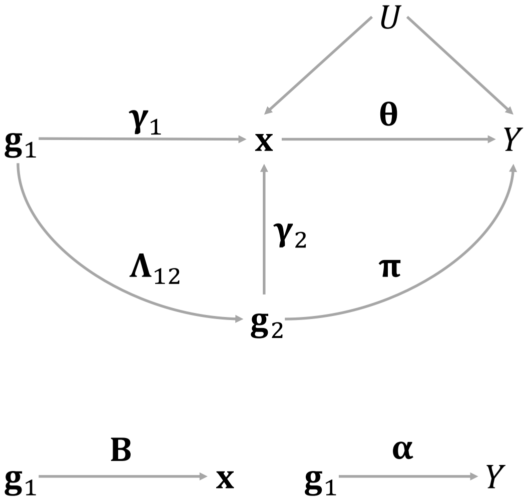

**Fig. 9** DAG representing the models specified in section 3.1. $\mathbf{g}_1 : m_1 \times 1$ is a vector of SNP genotypes used as IVs in MR to estimate $\boldsymbol{\theta}$, $\mathbf{g}_2 : m_2 \times 1$ is a generic vector of genotypes for other SNPs no in $\mathbf{g}_1$, $\mathbf{x} : p \times 1$ is a vector of expressions for $p$ genes in a tissue, $Y$ is the outcome trait, and $U$ is a generic confounding. If $\mathbf{g}_1$ and $\mathbf{g}_2$ are in LD, $\boldsymbol{\Lambda}_{12} \neq \mathbf{0}$. If $\mathbf{g}_2$ is associated with $Y$ conditional on $\mathbf{x}$, $\boldsymbol{\pi} \neq \mathbf{0}$.

In practice, we only have access to GWAS estimates of $(\boldsymbol{\alpha}, \mathbf{B})$, which we denote as $(\hat{\boldsymbol{\alpha}}, \hat{\mathbf{B}})$. Therefore, we use the following model to estimate $\boldsymbol{\theta}$:

$$\hat{\boldsymbol{\alpha}} = \hat{\mathbf{B}}\boldsymbol{\theta} + \boldsymbol{\Lambda}_{12}^{\top}\boldsymbol{\pi} + \tilde{\boldsymbol{\epsilon}}, \tag{8}$$

where $\tilde{\boldsymbol{\epsilon}}$ contains the measurement errors $\hat{\boldsymbol{\alpha}} - \boldsymbol{\alpha}$ and $\hat{\mathbf{B}} - \mathbf{B}$s. When estimating $\boldsymbol{\theta}$ in Equation 7 using only $\hat{\mathbf{B}}$, there will be horizontal pleiotropy bias if $\boldsymbol{\Lambda}_{12}^{\top}\boldsymbol{\pi} \neq \mathbf{0}$, which may be considered unbalanced if $m^{-1}\mathbf{1}_m^{\top}\boldsymbol{\Lambda}_{12}^{\top}\boldsymbol{\pi} \neq 0$ and correlated horizontal pleiotropy (CHP) if the correlation between $\boldsymbol{\Lambda}_{12}^{\top}\boldsymbol{\pi}$ and $\text{vec}(\hat{\mathbf{B}})$ is not $\mathbf{0}$. It was shown above that $\boldsymbol{\pi}$ is the association of $\mathbf{g}_2$ with $Y$ conditional on $\mathbf{g}_1$. Next we aim to provide an expression for the joint distribution of $(\hat{\mathbf{B}}, \boldsymbol{\Lambda}_{12}^{\top}\boldsymbol{\pi})$ to identify the potential sources of CHP bias in Equation 7.

First, we state the marginal distribution of $\hat{\mathbf{B}} = \mathbf{B} + \mathbf{W}_{\beta}$ where $\mathbf{B} = (\boldsymbol{\beta}_k)_{k=1}^{p}$ and $\mathbf{W}_{\beta}$ are random. As in [3], let

$$\boldsymbol{\beta}_k \sim \epsilon_k \mathrm{N}\left(\mathbf{0}, \frac{h_k^2}{\tilde{m}_k}\mathbf{I}_{m_1}\right) + (1 - \epsilon_k)\mathrm{N}(\mathbf{0}, \mathbf{0}) \tag{9}$$

be a mixture of $\tilde{m}_k$ SNPs that are associated with the expression of gene $k$ and $m_1 - \tilde{m}_k$ that are not. We specify this mixture explicitly because in the data it is true since the total set of $m$ SNPs used in MR is not a set of SNPs associated with the expression levels of *all* genes in a group and tissue, but typically only a subset of genes. The estimation error $\mathbf{W}_\beta = (\mathbf{w}_{\beta_k})_{k=1}^p$ is uncorrelated with $\mathbf{B}$ and has the distribution

$$\mathbf{w}_{\beta_k} \sim \mathrm{N}\left(\mathbf{0}, \frac{1}{n_k}\mathbf{R}_1\right) \tag{10}$$

for all $m_1$ SNPs where $n_k$ is the sample size in the GWAS for the expression of the $k$th gene. In MR, we have $\hat{\mathbf{B}} = (\hat{\boldsymbol{\beta}}_k)_{k=1}^p$, whose columns will have the distribution

$$\hat{\boldsymbol{\beta}}_k \sim \epsilon_k \mathrm{N}\left(\mathbf{0}, \left[\frac{h_k^2}{\tilde{m}_k}\mathbf{I} + \frac{1}{n_k}\mathbf{R}_1\right]\right) + (1-\epsilon_k)\mathrm{N}\left(\mathbf{0}, \frac{1}{n_k}\mathbf{R}_1\right). \tag{11}$$

Recall that CHP bias can arise when $\hat{\mathbf{B}}$ is correlated with $\boldsymbol{\Lambda}_{12}^\top \boldsymbol{\pi}$. Let $\boldsymbol{\Lambda}_{12}^\top(\boldsymbol{\tau}^\top \otimes \mathbf{R}_{12}^\top) := \mathrm{Cov}(\mathrm{vec}[\mathbf{B}], \boldsymbol{\Lambda}_{12}^\top \boldsymbol{\pi})$ where $\boldsymbol{\tau}^\top : p \times 1$ represents genetic covariance between $\boldsymbol{\pi}$ and the columns of $\mathbf{B}$. For example, consider $\mathrm{Cov}(\boldsymbol{\beta}_k, \boldsymbol{\pi}) := \tau_k \mathbf{R}_{12}^\top = [\mathrm{E}(\beta_{jk}\pi_s r_{js})]_{j,s=1}^{m_1,m_2}$. Since this covariance has a kronecker product structure, it can be zero if either one of two conditions are met, namely if (i) $\boldsymbol{\tau} = \mathbf{0}$ or (ii) $\mathbf{R}_{12}^\top = \mathbf{0}$. In principle, these conditions are met if either (i) the association between $\mathbf{g}_2$ and $Y$ *conditional on* $\mathbf{g}_1$ is independent of the *total* association between $\mathbf{g}_1$ and $Y$ or (ii) $\mathbf{g}_1$ and $\mathbf{g}_2$ are not in LD.

##### 2.1.3 CHP bias in traditional MR methods

Summary-based MR (SMR) [38] and MR-Robin [15], MR methods incorporating eQTLs that can only include a single gene in causal estimation, may suffer from UHP and/or CHP bias because of nonzero $\mathbf{R}_{12}$ and $\boldsymbol{\pi}$ for neighboring genes. In this section, we aim to better understand the extent to which SMR (a simmpler version of MR-Robin more popularly used) is vulnerable to UHP and CHP bias when considering Alzheimer's disease (AD) as the outcome trait and the expressions in blood of genes on chromosome (CHR) 19 using the real data that we used in the main text. First, we identified mutually exclusive groups of genes using the procedure described in **Methods**. First we define some notation within a group of genes. Let $\mathcal{M}$ denote the set of $M$ SNPs used as IVs for the entire group of $p$ genes, $\mathcal{M}_k$ denote the set of $m_k$ SNPs that are IVs for the $k$-th gene, $\mathbf{R}_k$ be the LD matrix for this gene, $\mathcal{M}_k^\perp$ be the set of SNPs in $\mathcal{M}$ but not in $\mathcal{M}_k$ that are in LD with SNPs in $\mathcal{M}_k$ via $\mathbf{R}_{k,-k}$, $\boldsymbol{\Lambda}_{k,-k} \approx \mathbf{R}_{k,-k}^\top \mathbf{R}_k^{-1}$, and $\boldsymbol{\pi}_{-k}$ be the association between SNPs in $\mathcal{M}_k^\perp$ and AD risk conditional on SNPs in $\mathcal{M}_k$. We estimated the following quantities:

$$I_1 = \|\mathbf{R}_k^{-1/2}\boldsymbol{\Lambda}_{k,-k}^\top \boldsymbol{\pi}_{-k}\|_2^2, \tag{12}$$

$$I_2 = \frac{1}{m_k}\mathbf{1}_{m_k}^\top \mathbf{R}_k^{-1/2}\boldsymbol{\Lambda}_{k,-k}^\top \boldsymbol{\pi}_{-k}, \tag{13}$$

$$I_3 = \theta_k - (\boldsymbol{\theta})_k, \tag{14}$$

where $\theta_k$ is estimated in univariable (single-gene) MR and $(\boldsymbol{\theta})_k$ is the multivariable (multiple-gene) MR estimate for the corresponding $k$th gene. Let $\hat{\boldsymbol{\delta}}_k := \hat{\boldsymbol{\alpha}}_k - \hat{\mathbf{B}}_k\hat{\theta}_k$. Below we list estimands for each of these quantities and their corresponding distributions under specified null hypotheses:

$$\hat{I}_1 = \|\boldsymbol{\Sigma}_{\Delta k}^{-1/2}\hat{\boldsymbol{\delta}}_k\|_2^2 \sim \chi^2(m_k) \quad \text{under} \quad H_0 : \boldsymbol{\Lambda}_{k,-k}^\top \boldsymbol{\pi}_{-k} = \mathbf{0} \tag{15}$$

$$\hat{I}_2 = \frac{1}{m_k}\mathbf{1}_{m_k}^\top \boldsymbol{\Sigma}_{\Delta k}^{-1/2}\hat{\boldsymbol{\delta}}_k \sim \mathrm{N}(\mathbf{0}, \eta) \quad \text{under} \quad H_0 : \frac{1}{m_k}\mathbf{1}_{m_k}^\top \boldsymbol{\Lambda}_{k,-k}^\top \boldsymbol{\pi}_{-k} = \mathbf{0} \tag{16}$$

$$\hat{I}_3 = \|\hat{\sigma}_\Theta^{-1/2}[\hat{\theta}_k - (\hat{\boldsymbol{\theta}})_k]\|_2^2 \sim \chi^2(1) \quad \text{under} \quad H_0 : \theta_k = (\boldsymbol{\theta})_k, \tag{17}$$

where

$$\eta := \frac{1}{m_k^2}\mathbf{1}_{m_k}^\top \boldsymbol{\Sigma}_\Delta \mathbf{1}_{m_k} \approx \frac{1}{m_k}, \tag{18}$$

$$\boldsymbol{\Sigma}_\Delta = \mathrm{Cov}(\hat{\boldsymbol{\delta}}_k) \tag{19}$$

$$\hat{\boldsymbol{\Sigma}}_\Delta = \mathbf{R}_k + \hat{\theta}_k^2 \sigma_{\mathbf{W}_\beta \mathbf{W}_\beta}^{2(k)} \mathbf{R}_k - 2\hat{\theta}_k \sigma_{\mathbf{W}_\beta \mathbf{w}_\alpha}^{(k)} \mathbf{R}_k \tag{20}$$

and

$$\hat{\sigma}_\Theta^2 = \widehat{\mathrm{Var}}(\hat{\theta}_k) + \widehat{\mathrm{Var}}[(\hat{\boldsymbol{\theta}})]_{k,k} - 2\widehat{\mathrm{Cov}}[\hat{\theta}_k|\hat{\boldsymbol{\beta}}_k, (\hat{\boldsymbol{\theta}})_k|\hat{\mathbf{B}}]. \tag{21}$$

Where $\hat{\sigma}_k$ and $(\hat{\sigma})_k$ represent the estimated standard deviations of the residuals during estimation of $\hat{\theta}_k$ and $\hat{\boldsymbol{\theta}}_k$ and $\mathbf{R}_{k(k)}$ is the LD matrix between valid IVs (see below for criteria) used in their respective estimators, $\hat{\sigma}_\Theta^2 = \mathbf{A}_k^{-1}\hat{\sigma}_k(\hat{\sigma})_k\mathbf{R}_{k(k)}\mathbf{A}_{(k)}^{-\top}$ for constant matrices $\mathbf{A}_k$ and $\mathbf{A}_{(k)}$. Regarding $I_2$, since $\hat{\boldsymbol{\delta}}_k$ is the estimated residual from linear regression by MR using the expression of gene $k$ as the exposure, $\hat{\boldsymbol{\delta}}$ is guaranteed to have a sample mean of $\mathbf{0}$. However, for each of $p$ genes in a group, we used the MRBEE estimator [21] with IMRP adjustment [37]. This method can estimate $\theta_k$ without bias from horizontal pleiotropy using a subset of the $m_k$ IVs, after which $\hat{\boldsymbol{\delta}}_k$ will become a reliable estimator for $\boldsymbol{\delta}_k$ (see []) and $\hat{\boldsymbol{\delta}}_k$ is not guaranteed to have a sample mean of $\mathbf{0}$.

To obtain an unbiased estimate for $\theta_k$, we also applied the following restrictions on the IV set: (i) P-value for association with gene expression less than 5x10$^{-5}$, (ii) absolute LD between SNPs used to instrument expression of the $k$th gene less than 0.9, (iii) $\geq$10 candidate IVs evaluated by MRBEE (some of which may have been further excluded due to evidence of nonzero horizontal pleiotropy at $P < 0.05$ using the tests in [21, 37]), and (iv) LD between the $j$th of $m_k$ SNPs and the $M - m_k$ other SNPs in the group less than 0.2. The latter worked to reduce bias from CHP via nonzero $\mathbf{R}_{k,-k}$ while still retaining enough SNPs for efficient estimation of $\hat{\theta}_k$. Regarding $I_3$, rejection of the corresponding null hypothesis is evidence of omitted-variable bias (OVB) (see [21]), which can be due to mediation or confounding (CHP). Both may be considered biased causal estimates, although where this bias is due to CHP or mediation cannot be determined by the test for $I_3 \neq 0$.

Under these restrictions to obtain a valid IV set, univariable MR using bias-corrected SMR (i.e., single-exposure MRBEE [21]) could only be performed for 194 of the 752 ( 25%) total genes with *cis*-eQTLs on CHR 19. This is another major limitation of univariable MR - the valid IV set can be reduced so small that causal estimation becomes unreliable and therefore should not even be performed. Figures 10, 11, 12, and 13 provide some inference for $I_1$, $I_2$, and $I_3$, respectively. These results indicate substantial nonzhero unbalanced horizontal pleiotropy across CHR 19. Subsequently, there is widespread evidence of differences in causal estimates made using univariable vs multivariable causal estimates, suggesting the presence off OV bias that may be due to CHP. We found that 37.6% of genes tested using univariable MR on CHR 19 (73/194) had evidence ($P < 5 \times 10^{-5}$) of nonzero horizontal pleiotropy ($I_1 \neq 0$), 13.7% of which had evidence of imbalance ($I_2 \neq 0$), and 48.2% of genes had multivariable causal estimates that differed from univariable causal estimates ($I_3 \neq 0$ where the test was available [see Figure 13 caption]). See the corresponding figure captions for more details.

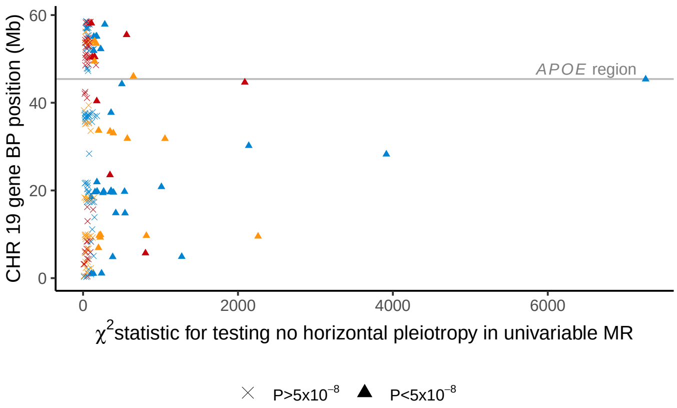

**Fig. 10** These results display $\chi^2$ test statistics for testing $H_0 : I_1 = 0$ as stated above. Each point represents a gene. This is a test for nonzero horizontal pleiotropy in the IV set in univariable MR using MRBEE [21], which can be considered a version of SMR [35, 38] corrected for bias from horizontal pleiotropy, weak instruments, sample overlap, and measurement/estimation error. This test was performed for each gene in CHR 19 that was put into a gene group in the main text. Different point colors represent distinct gene groups (see main text for how these groups were formed), with colors alternating from bottom to top on the y-axis from blue to red to yellow. Triangles represent genes for which $H_0$ is rejected at the level of genome-wide significance (i.e., $P < 5 \times 10^{-8}$); crosses represent genes for which $H_0$ is not rejected. The genomic region surrounding the *APOE* gene (known to be highly relevant for Alzheimer's disease risk) is labelled with a horizontal grey line. These results indicate substantial horizontal pleiotropy for many genes on this chromosome, where the strongest evidence of horizontal pleiotropy is observed in the *APOE* region. Only results for which univariable MR could be reliably performed are displayed (see text above figure).

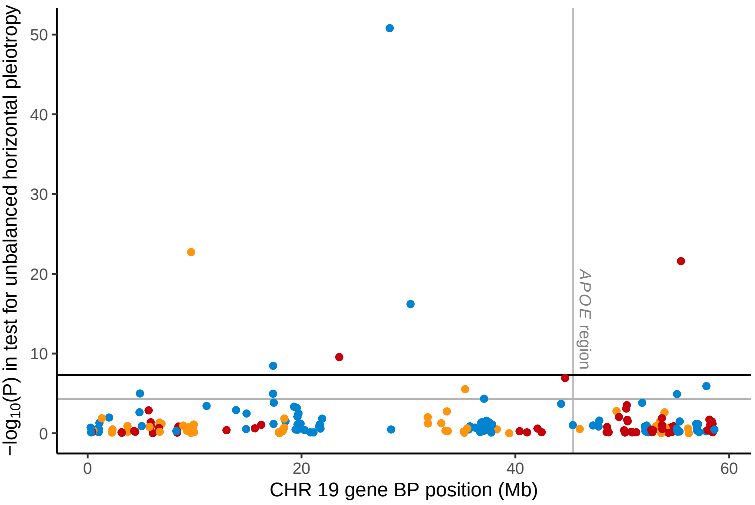

**Fig. 11** This is a test for unbalanced (i.e., nonzero mean) horizontal pleiotropy in univariable MR with MRBEE [21], which can be considered a version of SMR [35, 38] corrected for bias from horizontal pleiotropy, weak instruments, sample overlap, and measurement/estimation error. Each point represents a gene. This test was performed for each gene in CHR 19 that was present in a gene group in the main text. Genome-wide ($P < 5 \times 10^{-8}$) and marginal ($P < 5 \times 10^{-5}$) significance thresholds are displayed by black and gray horizontal lines, respectively. Different point colors represent distinct gene groups (see main text for how these groups were formed), with colors alternating from left to right on the x-axis from blue to red to yellow. The genomic region surrounding the *APOE* gene (known to be highly relevant for Alzheimer's disease risk) is labelled with a vertical grey line. These results indicate that many genes have evidence of unbalanced horizontal pleiotropy in univariable MR, including genes in the *APOE* region. Only results for which univariable MR could be reliably performed are displayed (see text above Figure 10).

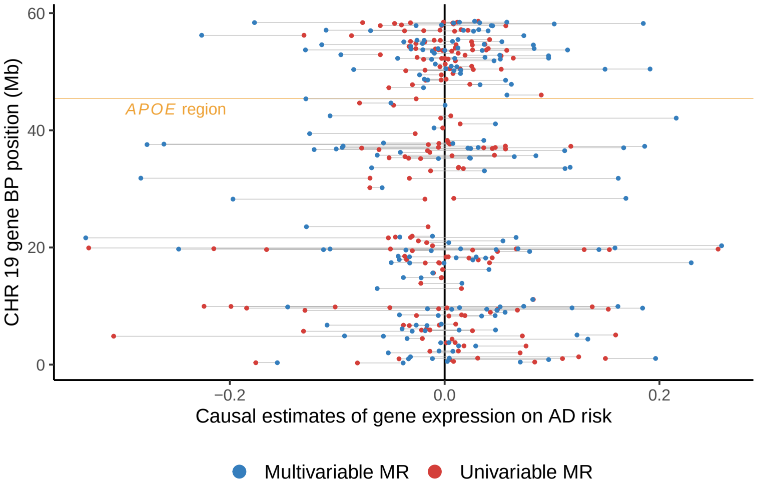

**Fig. 12** These results display differences between causal estimates made for each gene on chromosome (CHR) 19 using univariable MR vs multivariable MR. Each pair of points (paired by horizontal grey lines) corresponds to a single gene. Causal estimates were made using MRBEE [21], which can be considered a version of SMR [35, 38] corrected for bias from horizontal pleiotropy, weak instruments, sample overlap, and measurement/estimation error. Blue points represent multivariable MR estimates and red points represent univariable MR estimates. An absence of ommitted variable (OV) bias across CHR 19 would be observed if all red and blue points overlapped. Differences between these points for each gene, represented by horizontal grey lines, indicates OV bias, which is observed for many genes across CHR 19. The *APOE* gene region is highlighted by the yellow horizontal line, in which OV bias is observed. Only results for which univariable MR could be reliably performed are displayed (see text above Figure 10).

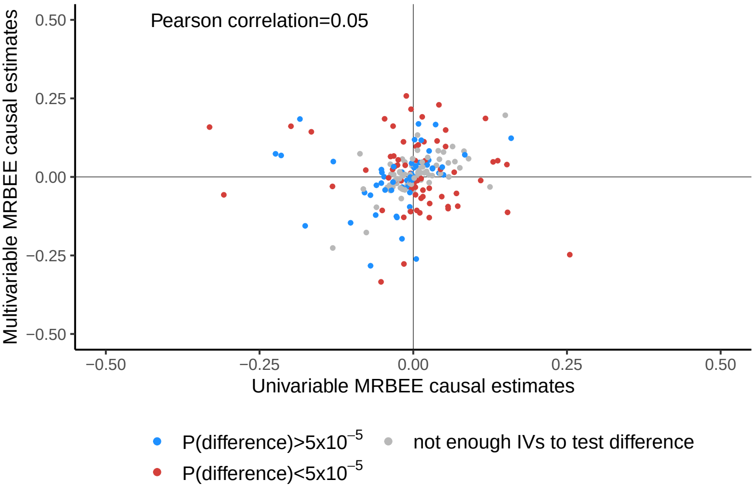

**Fig. 13** These results display the bivariate association between univariable MR and multivariable MR causal estimates and indications of the significance in testing $H_0 : I_3 = 0$ from the text above. Red and blue points respectively represent genes for which this null hypothesis was rejected and not rejected at P$< 5 \times 10^{-5}$. Grey points correspond to genes for which the test could not be reliably performed because of imprecise variance estimation in $\hat{I}_3$. For these genes, we could not estimate a positive $\hat{\sigma}^2_\Theta$ (see Equation 21) because of very small valid IV counts in univariable MR. 'Pearson correlation' corresponds to the linear correlation between univariable and multivariable MR causal estimates. This value will be 1 if there is no omitted variable (OV) bias (due either to CHP or mediation effects) and will approach 0 as OV bias becomes stronger.

##### 2.1.4 HORNET CHP correction

The method of protecting against CHP bias from eQTLs from other loci that are in LD with eQTLs in a target locus is outlined in Figure 14.

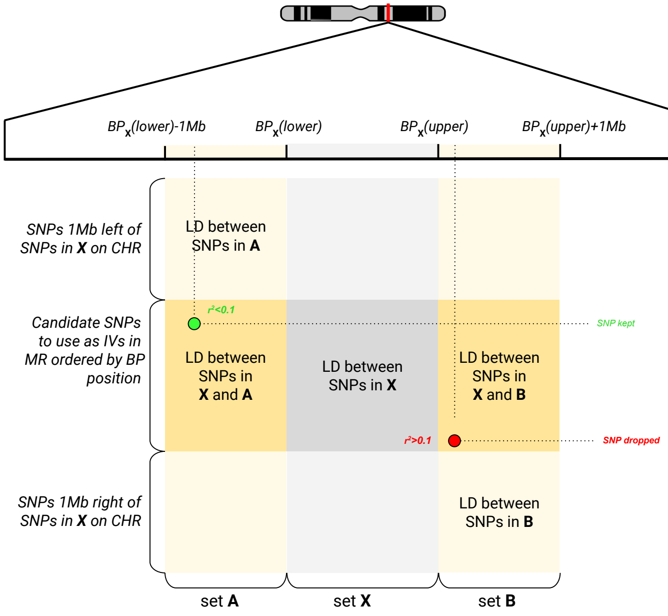

**Fig. 14** Visual depiction of how the final IV sets were pruned based on their LD with non-IVs within surrounding Mb windows of defined size (1Mb as the example in this figure). It was shown above that horizontal pleiotropy bias may be present in MR if the IVs used are in LD with SNPs that are conditionally associated with the outcome trait given the exposures. To reduce the potential for this bias, we subsetted the original IV set to only those SNPs that were not in LD $r^2 > 0.1$ with any SNPs within a 1Mb window outside of the BP range of the original IV set. This is displayed graphically as the green and red points in the figure, where $\mathbf{A}$ and $\mathbf{B}$ are sets of SNPs within 1Mb of the minimum and maximum BP positions of the original IV set, respectively.

#### 266 2.2 MRBEE bias-correction under LD

We now aim to demonstrate the validity of the MRBEE bias-correction in the case of correlated IVs (i.e., SNPs in LD used as IVs in MR). The original MRBEE theory [] was based on independent IVs, but we demonstrate here that the bias-correction in that case is the same as in our case of correlated IVs under a fixed-effects model for $\mathbf{B}$. Let $\hat{\mathbf{B}} = (\hat{\boldsymbol{\beta}}_j)_{j=1}^m = (\hat{\beta}_{jk})_{j=1,k=1}^{m,p}$ be the $m \times p$ matrix of GWAS estimates of the $m$ IVs on the expressions of $p$ genes in a tissue (MR exposures), $\hat{\boldsymbol{\alpha}}_j = (\hat{\alpha}_j)_{j=1}^m$ be the $m$-length vector of IV estimates of association with the outcome, and the IVs have the $m \times m$ positive definite LD correlation matrix $\mathbf{R} = (r_{js})_{js=1}^m$, where $\mathbf{P} = \mathbf{R}^{-1}$. We now assume a fixed effects model for $(\boldsymbol{\alpha}, \mathbf{B})$ for the purposes of causal estimation. This may be considered equivalent to causal estimation using MR conditional on the true causal SNPs used to instrument the exposures. We defined measurement error models for $(\hat{\boldsymbol{\beta}}_j, \hat{\alpha}_j)$ as in [21] in the following way:

$$\begin{pmatrix} \hat{\boldsymbol{\beta}}_j \\ \hat{\alpha}_j \end{pmatrix} = \begin{pmatrix} \boldsymbol{\beta}_j + \mathbf{w}_{\beta_j} \\ \alpha_j + w_{\alpha_j} \end{pmatrix} \sim N\left( \begin{bmatrix} \boldsymbol{\beta}_j \\ \boldsymbol{\alpha}_j \end{bmatrix}, \boldsymbol{\Lambda} := \begin{bmatrix} \boldsymbol{\Sigma}_{\mathbf{w}_\beta \mathbf{w}_\beta} & \boldsymbol{\sigma}_{\mathbf{w}_\beta w_\alpha} \\ \boldsymbol{\sigma}^\top_{\mathbf{w}_\beta w_\alpha} & \sigma^2_{w_\alpha} \end{bmatrix} \right), \tag{22}$$

where the errors in our measurements of $(\hat{\boldsymbol{\beta}}_j, \hat{\alpha}_j)$ were due only to sampling error introduced by finite GWAS sample sizes, and $(\boldsymbol{\beta}_j, \alpha_j)$ are fixed. MRBEE [20] makes a bias-correction to the IVW [5] estimating equation, which we denote as $S_{IVW}(\boldsymbol{\theta})$. Let $\mathbf{W}_\beta = (\mathbf{w}_{\beta_j})_{j=1}^m$ and $\mathbf{w}_\alpha = (w_{\alpha_j})_{j=1}^m$. It is shown in [21] that

$$\mathrm{E}[S_{IVW}(\boldsymbol{\theta})] = -\mathrm{E}(\mathbf{W}_\beta^\top \mathbf{P} \mathbf{W}_\beta) + \mathrm{E}(\mathbf{W}_\beta^\top \mathbf{P} \mathbf{w}_\alpha). \tag{23}$$

MRBEE subtracts from $S_{IVW}(\boldsymbol{\theta})$ the quantities in Equation 23 to produce an unbiased estimate of the causal parameter $\boldsymbol{\theta}$. Our goal now is to show that the quantity in Equation 23 is equal to $-\mathrm{E}(\mathbf{W}_\beta^\top \mathbf{W}_\beta) + \mathrm{E}(\mathbf{W}_\beta^\top \mathbf{w}_\alpha)$. Under the normality assumption in Equation 22,

$$\bar{\mathbf{W}} := (\mathbf{W}_\beta, \mathbf{w}_\alpha) \sim \mathrm{MatrixNormal}(\mathbf{0}_{m\times(p+1)}, \mathbf{R}, \boldsymbol{\Lambda}), \tag{24}$$

where $\mathbf{R}$ represents covariance between rows and $\boldsymbol{\Lambda}$ covariance between columns. By the positive-definiteness of $\mathbf{P} := \mathbf{R}^{-1}$,

$$\mathbf{P}^{1/2}\bar{\mathbf{W}} \sim \mathrm{MatrixNormal}(\mathbf{0}, \mathbf{I}_m, \boldsymbol{\Lambda}) \tag{25}$$

and

$$\bar{\mathbf{W}}^\top \mathbf{P} \bar{\mathbf{W}} \sim \mathrm{Wishart}(m, \boldsymbol{\Lambda}). \tag{26}$$

The proof for $\mathrm{E}(\mathbf{W}_\beta^\top \mathbf{P} \mathbf{W}_\beta)$ follows immediately from the properties of the Wishart distribution. Define the permutation matrix $\mathbf{C}_1 := (\mathbf{I}_{p\times p}, \mathbf{0}_{p\times 1})_{p\times(p+1)}$ such that

$$\mathbf{C}_1 \bar{\mathbf{W}}^\top \mathbf{P} \bar{\mathbf{W}} \mathbf{C}_1^\top = \mathbf{W}_\beta^\top \mathbf{P} \mathbf{W}_\beta. \tag{27}$$

It follows that

$$\mathrm{E}(\mathbf{C}_1 \bar{\mathbf{W}}^\top \mathbf{P} \bar{\mathbf{W}} \mathbf{C}_1^\top) = m\mathbf{C}_1 \boldsymbol{\Lambda} \mathbf{C}_1^\top = m\boldsymbol{\Sigma}_{\mathbf{W}_\beta \mathbf{W}_\beta}, \tag{28}$$

which is the desired result. For $\mathrm{E}(\mathbf{B}_\beta^\top \mathbf{P} \mathbf{w}_\alpha)$, we will show the proof element-wise also following the properties of the Wishart distribution. Consider the following:

$$\left[ (\mathbf{P}^{1/2}\mathbf{W}_\beta)^\top (\mathbf{P}^{1/2}\mathbf{w}_\alpha) \right]_k \sim \sigma_{\beta\alpha}^{[k]} \chi^2(m), \tag{29}$$

for $k = 1, ..., p$, which has expectation $m\sigma_{\beta\alpha}^{[k]}$ and since $m\boldsymbol{\sigma}_{\beta\alpha} = m(\sigma_{\beta\alpha}^{[k]})_{k=1}^{p}$ the result is proven.

#### 2.3 Heritability estimation

Assume the random model for $\boldsymbol{\beta}_k : M \times 1$ as in Equation 9 holds for associations between $M$ SNPs and the expression of gene $k$ in a tissue (here $M$ is the total number of SNPs tested for association with gene expression). Let $\tilde{m}_k \leq M$ denote the total number of SNPs causally related to the expression of this gene in tissue which has SNP heritability $h_k^2$. In our analyses, all association estimates were standardized by estimated standard error in GWAS such that $\hat{\boldsymbol{\zeta}}_k \approx \sqrt{n_k}\hat{\boldsymbol{\beta}}_k$ for GWAS sample size $n_k$ and $\hat{\boldsymbol{\zeta}}_k$ was the unit of analysis. Under the assumptions in model 9,

$$h_k^2 = \left[\frac{\mathrm{E}(\hat{\boldsymbol{\zeta}}_k^\top \hat{\boldsymbol{\zeta}}_k)}{M} - 1\right]\frac{\tilde{m}_k}{n_k}. \tag{30}$$

This is seen immediately from the following result

$$\mathrm{E}(\hat{\boldsymbol{\zeta}}_k^\top \hat{\boldsymbol{\zeta}}_k) = \mathrm{trace}\left(n_k\left[\frac{h_k^2}{\tilde{m}_k}\mathbf{I} + \frac{1}{n_k}\mathbf{R}\right]\right) \tag{31}$$

from the original model in Equation 11. A natural estimate of $h_k^2$ is

$$\hat{h}_k^2 = \left[\frac{\hat{\boldsymbol{\zeta}}_k^\top \hat{\boldsymbol{\zeta}}_k - \frac{1}{n_k}}{M} - 1\right]\frac{\hat{\tilde{m}}_k}{n_k} \tag{32}$$

where the $-1/n_k$ term is introduced as a measurement/estimation error bias-correction [21]. In practice, $\tilde{m}_k$ is rarely known and so must be estimated from the data. We estimated this quantity using a procedure similar to that used by PLINK [27] where we let $2\hat{\tilde{m}}_k$ be the number of SNPs of the total $M$ with association $P < 5 \times 10^{-5}$ for $\hat{\zeta}_{jk}$ that are independent ($\mathrm{r}^2 < 0.05$) of all other $M - 1$ SNPs for the gene group. We assume the factor 2 on $\tilde{m}_k$ consistent with results in [25, 26, 34] that *cis*-eQTLs explain approximately 1/3rd of the SNP heritability and *trans*-eQTLS explain the rest. Note that whether you assume a random or fixed effects model for $\boldsymbol{\beta}_k$, the result in Equation 30 is the same. To see this, let $\hat{\boldsymbol{\zeta}}_k \sim \mathrm{N}(\boldsymbol{\zeta}_k, n_k^{-1}\mathbf{R})$ and use the same technique as in Equation 31 then rearrange to arrive at the result in Equation 32.

#### 2.4 Source of bias in MRBEE from a misspecified LD matrix

In MR with gene expression as the exposure(s) of interest, we use eQTLs as instrumental variables (IVs). Standard methods of performing MR assume that these eQTLs will be independent of each other. However, there may only be very few (e.g., less than 5) IVs in a cis-region that are significant eQTLs and also independent of each other. If we only have, for example, 5 IVs to perform MR, there may be little power to detect causal effects. A more powerful approach would include IVs that are in LD with each other, assuming that a larger set of correlated IVs can explain more variance in the expression of a gene than a smaller set of independent IVs. Performing MR with $m$ correlated IVs requires estimating their LD matrix $\mathbf{R}$, which is usually accomplished in practice by using an external LD reference panel from approximately the same population, such as the 1000 Genomes reference panels [12]. It is well-known that the IVW estimator, equivalent to a generalized least squares estimator, will not be biased by misspecification of $\mathbf{R}$. That is, if in practice we use $\hat{\mathbf{R}} \neq \mathbf{R}$, the IVW estimator will not be biased because of it. The IVW estimator is generally biased from other sources as described above and in [21].

MRBEE makes a bias-correction to IVW for these other sources of bias which include measurement error/weak IVs and sample overlap. The MRBEE estimator with a set of $m$ IVs with no evidence of horizontal pleiotropy is

$$\hat{\boldsymbol{\theta}}_{\texttt{MRBEE}} = \left(\hat{\mathbf{B}}^\top \hat{\mathbf{R}}^{-1} \hat{\mathbf{B}} - m\boldsymbol{\Sigma}_{W_\beta W_\beta}\right)^{-1} \hat{\mathbf{B}}^\top \hat{\mathbf{R}}^{-1} \hat{\boldsymbol{\alpha}}. \tag{33}$$

In contrast to IVW, if each $\hat{\alpha}_j$ is standardized such that it represents the Z-statistic for association between the $j$th IV and the outcome trait, then $\hat{\boldsymbol{\alpha}} \sim \mathcal{N}(\boldsymbol{\alpha}, \mathbf{R})$. If $\hat{\mathbf{R}} = \mathbf{R}$, then MRBEE is not biased by $\hat{\mathbf{R}}$. This follows from Equation 24 under the assumption that $\texttt{Var}(\hat{\boldsymbol{\alpha}}) = \mathbf{R}$. However, if $\hat{\mathbf{R}} \neq \mathbf{R}$ then the bias-correction to IVW that MRBEE makes is not correct and therefore $\hat{\boldsymbol{\theta}}_{\texttt{MRBEE}}$ may be biased. This can be seen by the following. In Equation 23, it was stated that the bias in the IVW estimating equation is

$$-\texttt{E}(\mathbf{W}_\beta^\top \hat{\mathbf{R}}^{-1} \mathbf{W}_\beta)\boldsymbol{\theta} + \texttt{E}(\mathbf{W}_\beta^\top \hat{\mathbf{R}}^{-1} \mathbf{w}_\alpha) \tag{34}$$

which MRBEE assumes to be $-m(\boldsymbol{\Sigma}_{W_\beta W_\beta}\boldsymbol{\theta} - \boldsymbol{\sigma}_{W_\beta w_\alpha})$. If $\hat{\mathbf{R}} \neq \mathbf{R}$, then the bias in Equation 34 is more complex and MRBEE does not correctly adjust for it.

We now aim to investigate the extent to which MRBEE will be biased by a misspecified value of $\hat{\mathbf{R}}$. In this section, we consider a simple case in which $\hat{\mathbf{R}} = \xi\mathbf{R} + (1-\xi)\mathbf{I}$ for some constant $0 \leq \xi \leq 1$. In section 2.5, we consider more complex cases in which the size of the LD reference panel also varies. We performed simulations with generated GWAS summary statistics for 100 IVs using the following models

$$(\boldsymbol{\alpha}, \mathbf{B}) \sim \mathcal{N}\left(\mathbf{0}, \boldsymbol{\Sigma}, \frac{1}{4}\mathbf{R}\right) \tag{35}$$

$$\boldsymbol{\Sigma} = \mathbf{D}\begin{pmatrix} 1.0 & 0.2 & 0.2 \\ 0.2 & 1.0 & 0.5 \\ 0.2 & 0.5 & 1.0 \end{pmatrix}\mathbf{D} \tag{36}$$

$$\mathbf{D} = 0.1\mathbf{I}_3 \tag{37}$$

$$\mathbf{R} = \text{AR1}(0.5) \tag{38}$$

$$\widehat{\mathbf{B}} \sim \mathcal{N}\left(\mathbf{B}, \frac{1}{50}\boldsymbol{\Sigma}, \mathbf{R}\right) \tag{39}$$

$$\widehat{\mathbf{R}} = \mathrm{AR1}(\xi), \qquad \xi \in \{0.0, 0.1, ..., 0.8, 0.9\} \tag{40}$$

$$\hat{\boldsymbol{\theta}}_{\mathrm{IVW}} = \arg\min_{\boldsymbol{\theta}} -\frac{1}{2}\left(\boldsymbol{\alpha} - \hat{\mathbf{B}}^\top\boldsymbol{\theta}\right)^\top \widehat{\mathbf{R}}^{-1}\left(\boldsymbol{\alpha} - \hat{\mathbf{B}}^\top\boldsymbol{\theta}\right) \tag{41}$$

$$\hat{\boldsymbol{\theta}}_{\mathrm{MRBEE}} = \arg\min_{\boldsymbol{\theta}} -\frac{1}{2}\left(\boldsymbol{\alpha} - \hat{\mathbf{B}}^\top\boldsymbol{\theta}\right)^\top \widehat{\mathbf{R}}^{-1}\left(\boldsymbol{\alpha} - \hat{\mathbf{B}}^\top\boldsymbol{\theta}\right) - \boldsymbol{\theta}^\top\boldsymbol{\Sigma}_{W_\beta W_\beta}\boldsymbol{\theta}, \tag{42}$$

$$\tag{43}$$

where the constants 1/4 and 1/50 respectively represent minor allele frequency and the proportion of measurement error variance to the total signal variance. These simulation models implicitly assume no measurement error in the outcome associations $\boldsymbol{\alpha}$, which is irrelevant for our purpose here because neither IVW nor MRBEE will have any more or less bias as measurement error is added or removed from $\boldsymbol{\alpha}$. We performed 10,000 simulations for each scenario in which $\xi$ varied and the results are presented in Figure 15.

These results indicate that IVW is consistently biased irrespective of how close the working LD matrix $\hat{\mathbf{R}}$ is the the true LD matrix $\mathbf{R}$. On the other hand, MRBEE is unbiased when $\hat{\mathbf{R}} = \mathbf{R}$, but incurs a small upward bias when $\xi < rho = 0.5$ and a small downward bias when $\xi > \rho = 0.5$. Each of these biases are smaller than the bias incurred by IVW, expect when an extremely dense AR1(0.9) structure is assumed to fit data that were generated from AR1(0.5), which is unlikely to ever occur in practice. Interestingly, MRBEE is unbiased when the LD matrix is assumed to be equal to the identity matrix, although its variance in this setting is greater than in other settings when a denser LD structure is assumed.

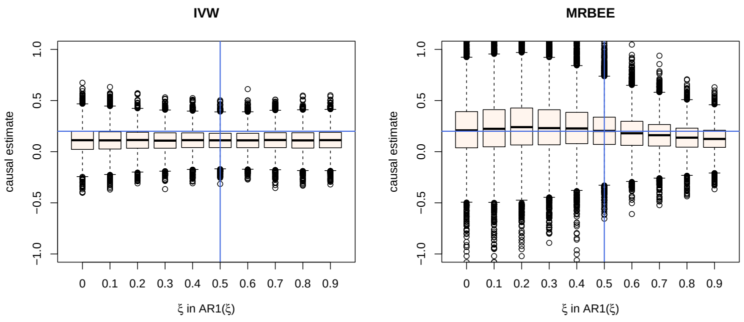

**Fig. 15** These are the results of 10,000 simulations, the settings for which are fully described in the text above. Displayed are boxplots of causal estimates made by IVW (left) and MRBEE (right) in different scenarios of assumed LD structure. The horizontal blue lines are positioned at the true causal effect, 0.2. The vertical blue lines are positioned at the value of $\rho$ which was used to generate the data.

#### 2.5 Misspecified LD

##### 2.5.1 Background

We mentioned in Section 2.4 that using a LD matrix in MR that is not equal to the true LD matrix representative of the discovery population can introduce bias in causal estimates using MRBEE. In this section, we demonstrate that misspecified LD of this type can cause inflation of test statistics corresponding to tests of the causal null hypothesis. MR methods that can allow for IVs that are in LD with each other are IVW [6], principal components analysis (PCA) [7], the conditional and joint (CoJo) algorithm [36], single-SNP [29, 31], LD pruning [11, 28], effective-median [19], and the JAM algorithm (joint analysis of marginal summary statistics) [23]. An estimate of the LD matrix between IVs is generally made using a reference panel and not the actual disease GWAS individual-level data. This is because reference panels are widely publicly available and individual-level data from many disease GWAS are not.

Using an independent reference panel to estimate LD between IVs used in MR may inflate test statistics and lead to a large false positive rate [19] if the reference population differs from the discovery (Figure 16) or if the reference panel is relatively small (Figure 3 in the main text). In the literature, only a single solution to this problem has been documented [19], but it is only available for univariable MR with gene expression, which may be highly vulnerable to bias and its own inflation because of complex regulatory networks between the expression levels of multiple nearby genes. Additionally, this correction relies on resampling methods that cannot be scaled genome-wide because of the computational burden. There is currently no solution to this problem of inflation from misspecified LD that can be applied to multivariable MR with gene expression.

We demonstrate that inflation in MR with gene expression is the result of relatively small reference panel sample sizes and systematic differences in genetic architecture between reference panel and discovery GWAS samples. Current methods with straightforward extensions to MVMR such as PCA and LD pruning are not guaranteed to control this inflation. We considered many potential solutions to this problem, the simulation results of which are presented in Sections 2.5.4 and 2.5.5.

##### 2.5.2 Inflation correction (IFC)

In the main text, we proposed a method to correct for inflation in MR test statistics due to misspecified LD structure amongst the IVs used in MR that is presented in Figure 17. Here, we describe that method in greater detail. We propose to correct for inflation by using a data-driven approach by using the degree of inflation in surrounding null regions to adjust the standard errors for causal estimates in the target region. Here, 'target region' refers to a locus in which there is a hypothesized causal relationship between the expression of one or more genes and the disease trait; 'null region' refers to a locus in which there is no evidence of any association between the genetic variants and the disease

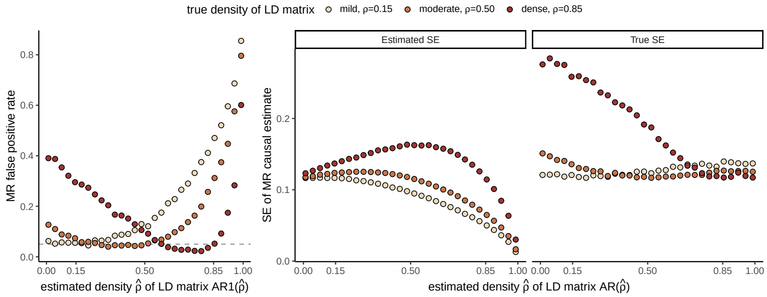

**Fig. 16** This figure demonstrates inflation in the Type I error rate of the IVW method [5] when applied to correlated IVs in simulation following a procedure similar to that described in Section 2.5.3. In this simulation, the size of the reference panel was fixed at 1,000 but the similarity in the true LD in the reference and discovery population varies. The true LD matrix $\mathbf{R}$ was of AR1($\rho$) structure and the true LD matrix in the reference panel was of AR1($\hat{\rho}$) structure. We display the false positive rate of IVW (left) for different value pairs of $\rho$ and $\hat{\rho}$, which are observed to surpass the nominal 0.05 level in many scenarios. We also display the IVW-estimated (middle) and corresponding true standard error (right) for the causal estimate in each of these scenarios. These results demonstrate that the estimated and true SEs are often unequal, which explains the inflation that is observed in the left panel.

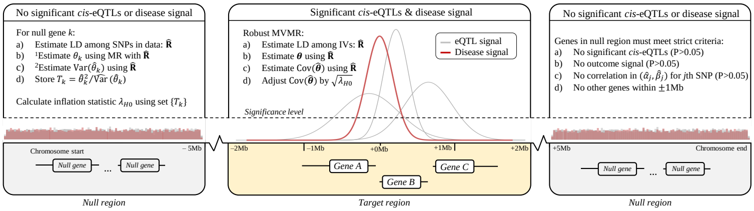

**Fig. 17** Visual description of the IFC method to correct for inflation in MVMR from misspecified LD. [1]: The causal effect $\theta_k$ can be estimated using any parametric multivariable MR method that allows the instrumental variables to be in LD. This should also be the same method that will later be used for inference in target gene regions. [2]: The variance of $\hat{\theta}_k$ should generally be estimated using robust methods, since it is unlikely to be true that the working LD matrix $\hat{\mathbf{R}}$ is exactly equal to the true LD matrix $\mathbf{R}_0$. This is because working LD matrices are typically estimated from reference samples of finite size and which may be ancestrally different from the discovery population.

trait or the expression of genes. In the truly null regions, the causal effects of gene expression on the outcome trait are each 0. This is because in Equation 7, all elements in $\boldsymbol{\alpha}$ and $\mathbf{B}$ are 0, implying that $\boldsymbol{\theta} = \mathbf{0}$ in the MR equation $\boldsymbol{\alpha} = \mathbf{B}\boldsymbol{\theta}$. By calculating inflation in these null regions, we are calculating inflation under $H_0 : \boldsymbol{\theta} = \mathbf{0}$. Any inflation that is observed in these regions is at least partially due to misspecified LD, and we assume that the same degree of inflation will be present in target regions. Under this assumption, we can adjust standard errors of causal estimates in target regions by the inflation observed in null regions. Figure 18 demonstrates that this assumption is reasonable using AD and eQTLs in blood tissue, evidenced by stable inflation across multiple null regions within chromosome 2 and across the entire genome.

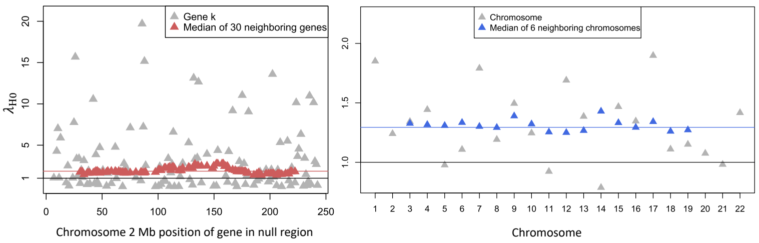

**Fig. 18** Left: observed inflation in null regions on chromosome 2. The horizontal black line is at the $\lambda_{H0} = 1$ position. The horizontal red line is at the position of the median of $\lambda_{H0}$ values across null regions on chromosome 2, which is 1.85. Right: observed inflation in null regions on all chromosomes. The horizontal black line is at the $\lambda_{H0} = 1$ position. The horizontal blue line is at the position of the median of $\lambda_{H0}$ across all chromosomes, which is at 1.29. These data are from eQTLs in blood tissue from [33] and AD GWAS summary statistics from [18].

Let $\hat{\theta}$ denote a causal estimate for the expression of a gene in a target region and $\lambda_{H0}$ denote the inflation observed in null regions. The corrected standard error estimate for $\hat{\theta}$ is the following:

$$\widehat{\mathrm{SE}}_\lambda(\hat{\theta}) = \widehat{\mathrm{SE}}(\hat{\theta})\sqrt{\lambda_{H0}}. \tag{44}$$

The obvious challenge lies in distinguishing truly null regions from those containing genes with extremely small causal effects. We therefore propose to use strict criteria for considering a genomic region as a null region. Firstly, it should be noted that we do not necessarily need to calculate inflation in null regions using multiple genes simultaneously in an MVMR framework. We may use one gene at a time to produce a set of causal estimate test statistics to be used in determining inflation. This is because under the condition that $\boldsymbol{\alpha} = \boldsymbol{\beta} = \mathbf{0}$ necessarily implies $\boldsymbol{\theta} = 0$, no negative confounding of $(\alpha_j, \beta_j)$ could exist to provide an alternative explanation for $\mathrm{E}(\hat{\theta}) \neq 0$. We therefore require that each gene to be used in determining inflation meet the following criteria: the SNPs within ±1Mb of the transcription start site are (i) not associated with the outcome, namely all P-values are greater than 0.05, (ii) not associated with the expression of the gene in the target tissue (all P>0.05), (iii) not within ±1Mb of any other genes, and (iv) the SNP associations with the outcome and gene expression are uncorrelated (P>0.05). In practice, conditions (i)-(iii) can be verified using the raw outcome phenotype and gene expression GWAS data. Condition (iv) can be verified by selected a set of SNPs for which *cis*-SNP association estimates are available for gene expression and the outcome, and calculating the empirical correlation. Generally, conditions (i)-(iv) should be satisfied after applying pruning to the raw LD matrix estimated by the reference panel. In our simulations below, we only consider SNPs that have LD coefficients less than 0.5 in absolute value.

##### 2.5.3 Simulation setup

In this section, we perform simulation to demonstrate the roles of $n_{\text{ref.}}$ and $\boldsymbol{\Psi}$ in inflating test statistics corresponding to causal effect estimates made using MVMR. These simulations used real data wherever possible. These data came from the Alzheimer's disease (AD) GWAS by [18] (n=455k) for the outcome trait and from the eQTLGen Consortium [33] (n=32k) for cis-eQTLs in blood tissue. We first identified a gene regulatory network in the 2q37.1 region that contained 7 genes and selected 484 candidate IVs for these genes using the following procedures. These IVs were jointly associated with the expression of at least one of the seven genes in blood tissue and were not in LD of $r^2$>0.1 with any other SNPs ±1Mb away from the network. We then estimated LD for these IVs using the 438k non-related European individuals in the UK Biobank [30] using the PLINKv1.9 software [27]. These data respectively provided the following quantities: $\hat{\boldsymbol{\alpha}} : 484 \times 1$, $\hat{\mathbf{B}} : 484 \times 7$, and $\widetilde{\mathbf{R}}_0 : 484 \times 484$. The values in $\hat{\boldsymbol{\alpha}}$ and $\hat{\mathbf{B}}$ were Z-scores, i.e., association estimates divided by their standard errors. The original LD matrix for the 484 SNPs estimated using UKBB was not positive definite. We applied LD pruning to $\widetilde{\mathbf{R}}_0$ using the threshold $|\tilde{r}_{ij}| < 0.85$ to generate the positive definite matrix $\mathbf{R}_0$. This subsetted our data from 484 to 168 SNPs.

From these data, we estimated genetic correlation between the columns of $\hat{\mathbf{B}}$ denoted as $\mathbf{S}$. We fixed heritability of gene expression at 0.05 [25] for each gene and at 0.01 for AD. We then perturbed the true LD matrix $\mathbf{R}_0$ and randomly drew it from a Wishart distribution to emulate real world conditions in which $\mathbf{R}_0$ is estimated from an external and sometimes relatively small reference panel. We did using the models:

$$\hat{\mathbf{R}} \sim \text{Wishart}(n_{\text{ref.}}, \mathbf{R}) \tag{45}$$

$$\mathbf{R} = \xi \mathbf{R}_0 + (1-\xi)\mathbf{I}_m \tag{46}$$

$$\xi \in \{0.0, 0.1, ..., 0.9, 1.0\} \tag{47}$$

$$n_{\text{ref.}} \in \{350, 450, ..., 950\}, \tag{48}$$

The minimum value in the set $n_{\text{ref.}}$ was chosen to be equal to the smallest population-specific sample size in 1000 Genomes Phase 3 [8], which corresponds to Hispanic individuals.

We therefore drew GWAS summary data for gene expression and AD from the following matrix normal distribution:

$$(\hat{\boldsymbol{\alpha}}, \hat{\mathbf{B}}) \sim \mathcal{N}(\mathbf{0}, \boldsymbol{\Sigma}, \hat{\mathbf{R}}), \qquad m \times (1+p) \tag{49}$$

where

$$\boldsymbol{\Sigma} = \mathbf{D}\begin{pmatrix} 1 & \boldsymbol{\sigma}_{\mathbf{x}Y}^{\top} \\ \boldsymbol{\sigma}_{\mathbf{x}Y} & \mathbf{S} \end{pmatrix}\mathbf{D}, \qquad (1+p)\times(1+p) \tag{50}$$

$$\mathbf{D} = \mathrm{diag}(0.01, 0.05, ..., 0.05), \qquad (1+p)\times(1+p) \tag{51}$$

and the quantity $\boldsymbol{\sigma}_{\mathbf{x}Y}$ was controlled to reflect the degree of causality between gene expressions $\mathbf{x}$ and AD $Y$. For example, $\boldsymbol{\sigma}_{\mathbf{x}Y} = \mathbf{0}$ implies no causality between $\mathbf{x}$ and $Y$ and was used to evaluate Type I error.

For each $(\xi, n_{\text{ref.}})$ pair, we drew $(\hat{\boldsymbol{\alpha}}, \hat{\mathbf{B}})$, applied the causal estimation methods of PCA [7], LD pruning [11], single SNP [29, 31], and our proposed IFC and recorded power and Type I error. For our IFC method, we require additional data beyond that which is provided by the 484 IVs. These data were the remaining gene expression and AD GWAS summary data on chromosome 2 that met the criteria for null regions as described in Section 2.5.2. We estimated Type I error when $\boldsymbol{\sigma}_{\mathbf{x}Y} = \mathbf{0}$ and power when $\sigma_{\mathbf{x}Y} = (\rho\sqrt{0.01 \times 0.05})$ where $\rho \in \{0.1, 0.2, 0.3\}$.

##### 2.5.4 Type I error

The results of these simulations suggest that the IVW [6] method has inflated Type I error when the true LD in the reference panel is sparser than that in the discovery population. IVW also has deflated Type I error when the reference and discovery populations have the same LD structure but the size of the reference panel is less than 1,000 individuals. Pruning at the $|r| < 0.5$ level reduced some of the Type I error inflation and deflation that was present in IVW, but did not bring Type I error to nominal levels (i.e., 0.05) in all simulation scenarios. Pruning at the $|r| < 0.3$ level removed the Type I error deflation, but not the inflation. The PCA method [7] had drastically inflated Type I error rates in all simulation scenarios. Using pruning at the $|r| < 0.3$ level then applying IFC controlled Type I error better than any other method or combination of methods and did not deflate Type I error below the nominal 0.05 level. This approach only had inflation of Type I error when the true LD matrix in the reference panel was much more dense than the true LD matrix in the discovery population, a situation which is unlikely to occur in practice. Importantly, pruning + IFC also controls Type I error when the size of the LD reference panel is small. Jackknifing methods generally still had inflated Type I error, though to a lesser extent than IVW, pruning alone, or PCA.

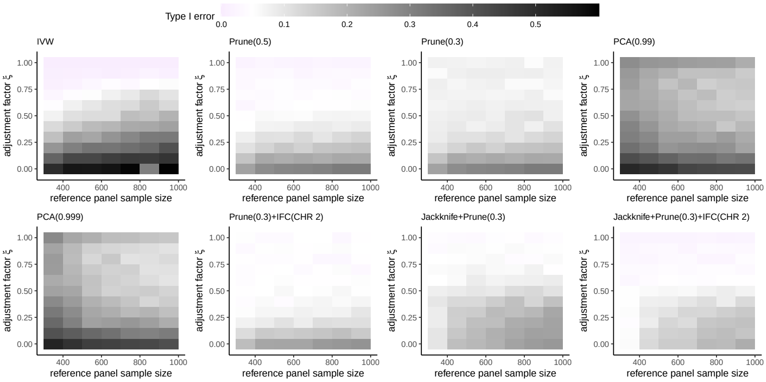

**Fig. 19** These results display Type I error for different methods for performing MR with instrumental variables that are in LD with each other. Full simulation settings are described in the text. Type I error is displayed for the first of 3 exposures. The PCA methods use univariable MR with only the first exposure; the other methods use multivariable MR with all three.

##### 2.5.5 Power

We also investigated power of each method when $\boldsymbol{\sigma}_{\mathbf{x}Y} = \boldsymbol{\theta} = (0.1)$ the results of which are displayed in Figure 20. These results demonstrate that the IVW method is generally most powerful at very specific combinations of the adjustment factor $\xi$ and reference panel sample size (see Figure 20). The power of IWV is low even when the true LD structure in the reference panel is the same as in the discovery population (i.e., $\xi = 1$), but the size of the reference panel is less than 1k. Only as the size of the reference panel increases can the IVW method achieve greater power when $\xi = 1$. Pruning at the $|r| < 0.5$ the $|r| < 0.3$ thresholds have similar power which increases as $\xi$ approaches 1 and the reference panel sample size increases. These methods can achieve greater power than IVW when the reference panel is relatively small. PCA methods actually have lower power as the reference panel sample size increases, and greater power as LD in the reference panel approaches the identity matrix, i.e. $\xi$ approaches 0. Our pruning and IFC corrective method generally has power than increases with $\xi$ approaching 1 and the reference panel sample size increasing. This approach generally has less power than alternative methods, which is the sacrifice made for controlling Type I error. Jackknife methods can generally be more powerful than all methods except pruning at $|r| < 0.5$ when LD in the reference panel is sparser than LD in the discovery population (i.e., $\xi < 0.5$). Overall, these results confirm that our corrective method of pruning and IFC does not sacrifice substantial power to achieve controlled Type I error.

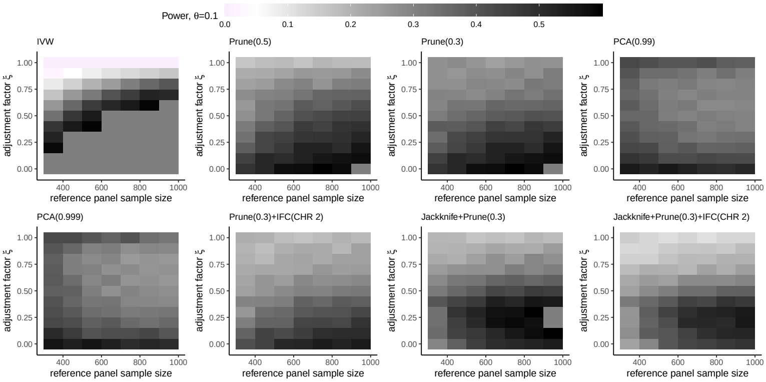

**Fig. 20** These results display power for different methods for performing MR with instrumental variables that are in LD with each other and $\theta = 0.1$. Full simulation settings are described in the text. Power is displayed for the first of three exposures. The PCA methods use univariable MR with only the first exposure; the other methods use multivariable MR with all three.

### 3 Estimating bias-correction terms

GScreen is an extension of the MR with unbiased estimating equations (MRBEE) method [21] to the high dimensional setting. MRBEE corrects for bias from weak instruments [1] that is introduced by measurement error in the GWAS associations. Let $(\hat{\alpha}_j, \hat{\beta}_{j1}, \hat{\beta}_{j2}, ..., \hat{\beta}_{jp}) = (\hat{\alpha}_j, \hat{\boldsymbol{\beta}}_j^\top)$ be a pair of associations between the $j$th IV and the outcome and expression of $p$ genes, respectively. It was shown in [21] that when estimating $\boldsymbol{\theta}$ from

$$\hat{\alpha}_j = \hat{\boldsymbol{\beta}}_j^\top \boldsymbol{\theta} + \varepsilon_j \tag{52}$$

using the standard IVW method [5], there is downward bias due to nonzero variance of $\boldsymbol{\beta}_j - \hat{\boldsymbol{\beta}}_j$, which we denote as $\boldsymbol{\Sigma}_{W_\beta W_\beta}$. MRBEE estimates $\boldsymbol{\Sigma}_{W_\beta W_\beta}$, denoted as $\widehat{\boldsymbol{\Sigma}}_{W_\beta W_\beta}$, to correct for bias in IVW [5]. The estimate $\widehat{\boldsymbol{\Sigma}}_{W_\beta W_\beta}$ is highly precise when there are many SNPs available that have no evidence of association with the expression of any of the $p$ genes in the locus. However, when there are relatively few SNPs available that meet this criteria for any pair of genes, the corresponding estimate in $\widehat{\boldsymbol{\Sigma}}_{W_\beta W_\beta}$ may be imprecise. When the total number of SNPs available to estimate $\boldsymbol{\Sigma}_{W_\beta W_\beta}$ is less than 50, HORNET automatically treats the corresponding elements in $\widehat{\boldsymbol{\Sigma}}_{W_\beta W_\beta}$ as missing and employs the maximum determinant method (MaxDet; [14]) to impute the missing values. In practice, $\widehat{\boldsymbol{\Sigma}}_{W_\beta W_\beta}$ is converted to its corresponding correlation matrix before MaxDet is applied.

Consider that $\widehat{\boldsymbol{\Sigma}}_{W_\beta W_\beta}$ is a $p \times p$ correlation matrix with 2 missing values corresponding to the correlation between measurement errors for a single pair of genes. MaxDet estimates the missing value in $\widehat{\boldsymbol{\Sigma}}_{W_\beta W_\beta}$ as that which maximizes the determinant of $\widehat{\boldsymbol{\Sigma}}_{W_\beta W_\beta}$ while retaining positive definiteness. The method is essentially an imputation procedure for the correlation matrix. In the real data, there may be missing measurement error correlation estimates for $k$ pairs of genes. In this case, HORNET applies MaxDet to each of $k$ sub-matrices of $\widehat{\boldsymbol{\Sigma}}_{W_\beta W_\beta}$ that contains only non-missing and non-imputed values.

### 4 Gene Selection

The HORNET software estimates causal effects of gene expression using a two-step procedure. First, genes with evidence of causality are identified using a screening tool called GScreen. Second, causal estimates of the selected genes are made using MRBEE [21].

G-Screen aims to perform gene selection while remaining robust to horizontal pleiotropy by using Huber weights to approximate quantile regression [17] and a SCAD [13] penalty to perform gene selection. Quantile regression is well-known to be robust in the presence of horizontal pleiotropy [2], which effectively indicates the presence of relatively large outliers in multiple regression, and the SCAD penalty will automatically exclude some genes from causal estimation, thereby selecting others. This procedure also includes the MRBEE bias-correction to prevent incorrect inference from weak instrument/measurement error biases during gene selection. Algorithm 1 summarizes this procedure. This algorithm requires the tuning parameters $\gamma$ and $\lambda$, which respectively control the degrees of penalization of horizontal pleiotropy and the number of genes selected. To choose an optimal $(\gamma, \lambda)$ pair, we search a grid of values and calculate the following BIC:

$$\text{BIC}_\gamma = \sum_j \xi(\varepsilon_j, \gamma)\log(\sigma_\xi^2) + \Big[\log(m - |\mathcal{S}_\eta|) + \log(p)\Big]|\mathcal{S}_\eta| \tag{53}$$

where $\sigma_\xi^2$ is the sum of squared and Huber-weighted residuals, $|\mathcal{S}_\eta|$ is the number of selected genes, $\xi(\varepsilon_j, \gamma)$ is the normalized Huber weight for the $j$th SNP (normalized to the range 0-1), $m$ is the number of SNPs, and $p$ is the total number of genes under study in a locus. In practice, weights $(w_j)$ are continually normalized such that the maximum value is always 1.

Figure 21 shows the performance G-Screen in screening genes for evidence of causality using eQTL GWAS data from the cortex [10] and AD GWAS data from [18] as an example. Of the 441 genes selected by G-Screen during screening, 49.7% were prioritized. Here, 'prioritized' refers to the condition when the gene has a corresponding causal effect P-value less than $5 \times 10^{-5}$ and its Pratt index value is larger than 0.1.

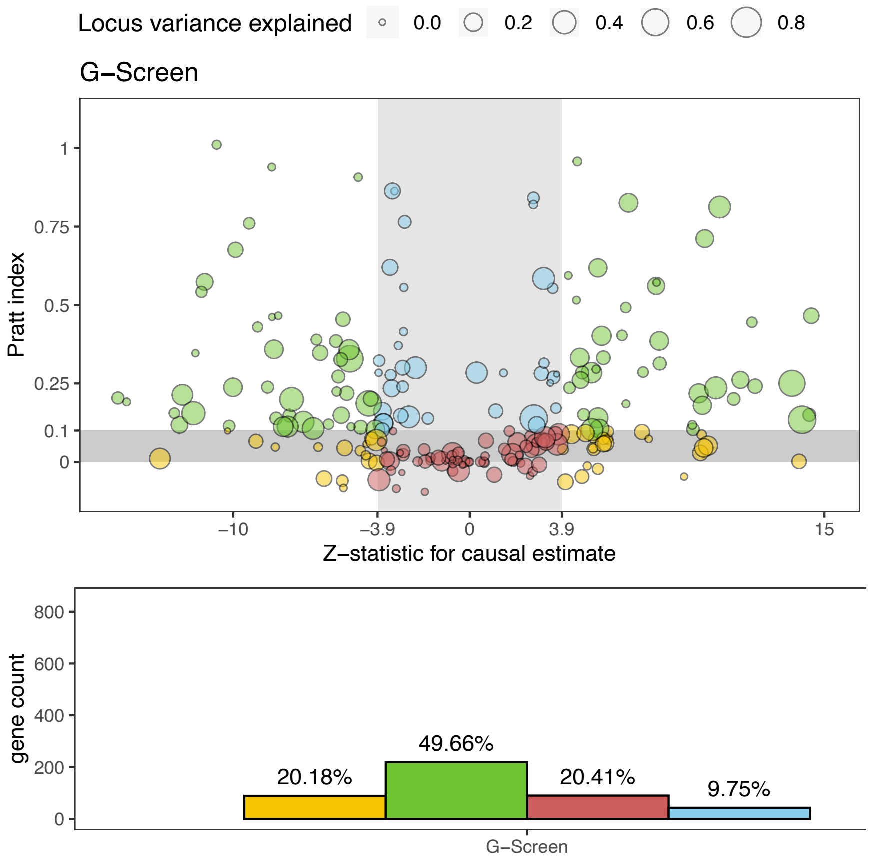

**Fig. 21** Displayed in the top panel is a volcano plot for HORNET analyses that were completed using eQTL GWAS data from cortex tissue [10] and Alzheimer's disease (AD) GWAS data from [18]. The x-axes represent the test statistics for testing the null hypothesis that a given gene does not have a causal effect on AD risk when expressed in the cortex. The y-axes represent Pratt index values. Each point in these figures corresponds to a single gene that was selected by G-Screen. The gray shaded ares are those in which the gene is not prioritized (i.e., the Z-statistic *and* Pratt index values are each not of sufficient magnitude. Prioritized genes are therefore those highlighted in green. The bottom panel displays counts and proportions of each classification of genes that are presented in the volcano plots in the top panel.

### 5 Prioritizing tissues

In this section, we describe how to use the `tissue_chooser.py` command-line tool to identify tissues in which a pre-defined candidate set of genes have the strongest eQTL signals. From GTEx data of 54 tissues [9], we estimated heritability scores for the expression of each available gene in each tissue using all significant cis-eQTLs. These heritability scores were calculated from cis

**Algorithm 1** Pseudo-code of G-Screen

**Require:** $m \times p$ eQTL-MVMR design matrix $\hat{\mathbf{B}}_0$ of SNP-gene expression associations for genes in set $\mathcal{S}$, $m \times 1$ vector of corresponding SNP associations with the disease phenotype $\hat{\boldsymbol{\alpha}}_0$, $m \times m$ inverse LD matrix $\boldsymbol{\Omega}$ between SNPs, MRBEE bias-correction terms $\widehat{\boldsymbol{\Sigma}}_{W_\beta W_\beta}$, Huber weight tuning parameter $\gamma$ for UHP/CHP in $\xi(\cdot, \gamma)$, SCAD tuning parameter $\lambda$ for gene selection, initial causal estimates $\hat{\boldsymbol{\eta}}^0$, tolerance $\epsilon$

**Transform**: $\hat{\mathbf{B}} = \boldsymbol{\Omega}^{1/2}\hat{\mathbf{B}}_0$, $\hat{\boldsymbol{\alpha}} = \boldsymbol{\Theta}^{1/2}\hat{\boldsymbol{\alpha}}_0$

**while** $\|\hat{\boldsymbol{\eta}}^{(t+1)} - \hat{\boldsymbol{\eta}}^{(t)}\|_2 > \epsilon$ **do**

Determine weights: $w_j = \xi(\hat{\alpha}_j - \hat{\boldsymbol{\beta}}_j^\top \hat{\boldsymbol{\eta}}^{(t)}, \gamma)$

Define $m_w = \sum_j w_j$, $\mathbf{D} = \mathrm{diag}(w_j)_{j=1}^m$

Update $\hat{\boldsymbol{\theta}}^{(t+1)} = \left(\hat{\mathbf{B}}^\top \mathbf{D}\hat{\mathbf{B}} - m_w \boldsymbol{\Sigma}_{W_\beta W_\beta} + \rho \mathbf{I}\right)^{-1}\left(\hat{\mathbf{B}}^\top \mathbf{D}\hat{\mathbf{B}} + \rho \mathbf{I} - \boldsymbol{\mu}^{(t)}\right)$

Penalize $\hat{\eta}_j^{(t+1)} = S_\lambda(\hat{\theta}_j^{(t+1)})$

Update $\mu^{(t+1)} = \boldsymbol{\mu}^{(t)} + \rho(\hat{\boldsymbol{\theta}}^{(t+1)} - \hat{\boldsymbol{\eta}}^{(t+1)})$

Find $\epsilon = \|\hat{\boldsymbol{\eta}}^{(t+1)} - \hat{\boldsymbol{\eta}}^{(t)}\|_2^2$

**end while**

**Ensure:** Set $\mathcal{S}_\eta = \{k : \hat{\eta}_k \neq 0 :\}$ for $k = 1, ..., p$ of genes with evidence of causality

SNPs within ±1Mb and are proportional to SNP heritability. We then created a matrix in which each row was a gene, each column was a tissue, and each value was a heritability score. The `tissue_chooser.py` tool simply receives a comma-separated list of gene IDs, or a header-less file in which each row is a gene ID, and returns the tissues for which the eQTLs in GTEx v8 are the strongest. We first show how heritability scores were calculated, then demonstrate how to use the tool from the command line.

#### 5.1 Heritability scores

Heritability scores are calculated in the following way. First, GTEx v8 associations between SNPs and gene expression in each tissue were calculated using fastQTL [24]. All associations that were significant at a corrected threshold were recorded and placed into the `GTEx_Analysis_v8_eQTL.tar` file available at https://gtexportal.org/home/datasets [9]. These data provided us with Z-statistics for association between the $k$ SNP and the $G$th gene in the $\mathcal{T}$th tissue, denoted here as $z_k^G(\mathcal{T})$. From these association estimates, we created the vectors $\mathbf{z}^G(\mathcal{T}) = [z_k^G(\mathcal{T})]_{k=1}^{p_G}$ of varying length $p_G$ that were gene-specific. In other words, these vectors contained all SNP-gene association estimates that were significant at a specific threshold in a specific tissue, and we created them for each gene-tissue pair. It was shown in [39] that $\mathrm{Cov}[\mathbf{z}^G(\mathcal{T})] = \mathbf{R}$, which is the LD matrix between the SNPs whose associations with gene expression are stored in $\mathbf{z}^G(\mathcal{T})$.

We calculated heritability scores as

$$H_S(G,\mathcal{T}) = \Big[\mathbf{z}^G(\mathcal{T})\Big]^\top \widehat{\mathbf{R}}^{-1}\Big[\mathbf{z}^G(\mathcal{T})\Big], \tag{54}$$

where $\widehat{\mathbf{R}}$ is an estimate of the corresponding LD matrix between the SNPs whose association estimates are in $\mathbf{z}^G(\mathcal{T})$, which we made using the full 1000 Genomes Phase 3 reference panel [8]. The heritability scores $H_S(G,\mathcal{T})$ were previously shown to be proportional to SNP heritability [4]. Since the tissue prioritizing tool that we present in this section is only intended to provide a ranked list of tissues in which the strongest eQTL signals for a pre-specified list of genes are, the heritability scores $H_S(G,\mathcal{T})$ are sufficient for accomplishing this task. Heritability scores for each gene-tissue pair were then stored in the file `hscores.txt`, which can be found in the `tissue_priority` directory of the HORNET software (see https://github.com/noahlorinczcomi/HORNET).

#### 5.2 Running `tissue_chooser.py` to prioritize tissues

In this subsection, we demonstrated how to use our `tissue_chooser.py` tool to automatically search `hscores.txt` for tissues with the strongest eQTLs. Please see the 'Choosing tissues' branch at https://github.com/noahlorinczcomi/HORNET for a complete demonstration of how to use this tool. Briefly, this tool receives either a comma-separated list or file of gene IDs to its `--candidateGenes` flag, a 'yes' or 'no' to its `--saveAsFile` flag indicating if the results should be saved in a file in addition to being printed to the console, and the output filepath to `--outFile` if you put 'yes' to the `--candidateGenes` flag. Note, the top results will always be printed to the console, unless you want to suppress them by setting the `--printResults` flag to be 'no'. An example of output that could be generated by this tool for the *HMGCR*, *CETP*, and *FES* genes is displayed in Figure 22.

```
The top 10 tissues for these genes are the following:

                                                          Genes  GeneCount nSignifSNPs   Maxh2Score
Tissue
Lung                            ENSG00000087237,ENSG00000182511          2       16,35  1208.473765
Cells_Cultured_fibroblasts      ENSG00000087237,ENSG00000182511          2        6,23  2949.393100
Muscle_Skeletal                                 ENSG00000113161          1          76  1548.315195
Whole_Blood                                     ENSG00000182511          1          25   837.521937
Thyroid                                         ENSG00000182511          1          19  1983.885576
Pancreas                                        ENSG00000182511          1          18  1139.772883
Heart_Atrial_Appendage                          ENSG00000087237          1           9   230.349358
Small_Intestine_Terminal_Ileum                  ENSG00000087237          1           9   227.685715
Stomach                                         ENSG00000087237          1           8   184.174806
Adipose_Visceral_Omentum                        ENSG00000113161          1           3    28.204502
```

**Fig. 22** This is the output of running the

`python tissue_chooser.py --candidateGenes ENSG00000113161,ENSG00000087237,ENSG00000182511`

command in the `HORNET` directory. All values are aggregated within tissues. `Maxh2score` is the maximum heritability score for the tissue. `nSignifSNPs` is the number of SNPs significantly associated with gene expression after adjustment for multiple comparisons in GTEx v8 [9]. `TissueCount` represents the number of genes for which the specific tissue is in the top 5 tissues with the strongest cis-eQTLs. `Genes` represents the genes for which each tissue contains one of the top 5 strongest eQTLs.

#### 5.3 Limitations

This tool has the following limitations. First, this tool relies solely on GTEx v8 data for the inferences it intends to supply. Second, these heritability scores are proportional to SNP heritability but are also proportional to the true total number of causal SNPs, which in this case may not be reliably estimated from only the cis-eQTL data. The tool therefore implicitly assumes constant numbers of causal SNPs across all genes in all tissues. In this context, 'causal SNPs' refers to those SNPs that cause variation in gene expression in a specific tissue. Thirdly, any prioritization of certain tissues over others is completely agnostic to the outcome phenotype. It therefore may be true that a gene with a very strong causal effect on a disease when expressed in one particular tissue may not have strong enough eQTLs in that tissue to give it a relatively high ranking by our `tissue_chooser.py` tool. Researchers should therefore only consult this tool as one of many forms of guidance in choosing the most appropriate tissues for their analysis. Fifthly, we used strictly GTEx summary data [9] when constructing the reference data set `hscores.txt` on which the `tissue_chooser.py` relies. The GTEx v8 sample size for whole blood tissue is 670, whereas the sample size for cis-eQTLs in the eQTLGen Consortium [33] is 31k, which provides more statistical power for detecting cis-eQTLs than GTEx v8. Since whole blood tissue is generally considered in all analyses anyway, we omitted calculation of heritability scores using eQTL GWAS in whole blood from the eQTLGen Consortium.

### 6 Software

The HORNET software is available as a command line tool and desktop application for Linux, Windows, and Mac machines. Complete tutorials demonstrating how to download and use the HORNET software are present at https://github.com/noahlorinczcomi/HORNET under the `README.md` and `HORNET_Desktop.md` files for the command line and desktop versions, respectively.

[3] Brendan Bulik-Sullivan, Hilary K Finucane, Verneri Anttila, Alexander Gusev, Felix R Day, Po-Ru Loh, Laramie Duncan, John RB Perry, Nick

Patterson, Elise B Robinson, et al. An atlas of genetic correlations across human diseases and traits. *Nat. Genet.*, 47(11):1236–1241, 2015.

[4] Brendan K Bulik-Sullivan, Po-Ru Loh, Hilary K Finucane, Stephan Ripke, Jian Yang, Nick Patterson, Mark J Daly, Alkes L Price, and Benjamin M Neale. Ld score regression distinguishes confounding from polygenicity in genome-wide association studies. *Nat. Genet.*, 47(3):291–295, 2015.

[5] Stephen Burgess and Jack Bowden. Integrating summarized data from multiple genetic variants in mendelian randomization: bias and coverage properties of inverse-variance weighted methods. *arXiv preprint arXiv:1512.04486*, 2015.

[12] Susan Fairley, Ernesto Lowy-Gallego, Emily Perry, and Paul Flicek. The international genome sample resource (igsr) collection of open human genomic variation resources. *Nucleic Acids Research*, 48(D1):D941–D947, 2020.

[13] Jianqing Fan and Runze Li. Variable selection via nonconcave penalized likelihood and its oracle properties. *Journal of the American statistical Association*, 96(456):1348–1360, 2001.

[14] Dan I Georgescu, Nicholas J Higham, and Gareth W Peters. Explicit solutions to correlation matrix completion problems, with an application to risk management and insurance. *Royal Society open science*, 5(3): 172348, 2018.

[15] Kevin J Gleason, Fan Yang, and Lin S Chen. A robust two-sample mendelian randomization method integrating gwas with multi-tissue eqtl summary statistics. *bioRxiv*, pages 2020–06, 2020.

[16] Trevor Hastie, Rahul Mazumder, Jason D Lee, and Reza Zadeh. Matrix completion and low-rank svd via fast alternating least squares. *The Journal of Machine Learning Research*, 16(1):3367–3402, 2015.

[17] Xuming He, Xiaoou Pan, Kean Ming Tan, and Wen-Xin Zhou. Smoothed quantile regression with large-scale inference. *Journal of Econometrics*, 2021.

[18] Iris E Jansen, Jeanne E Savage, Kyoko Watanabe, Julien Bryois, Dylan M Williams, Stacy Steinberg, Julia Sealock, Ida K Karlsson, Sara Hägg, Lavinia Athanasiu, et al. Genome-wide meta-analysis identifies new loci and functional pathways influencing alzheimer's disease risk. *Nature genetics*, 51(3):404–413, 2019.

[19] Lin Jiang, Lin Miao, Guorong Yi, Xiangyi Li, Chao Xue, Mulin Jun Li, Hailiang Huang, and Miaoxin Li. Powerful and robust inference of complex phenotypes' causal genes with dependent expression quantitative loci by a median-based mendelian randomization. *The American Journal of Human Genetics*, 109(5):838–856, 2022.

[20] Noah Lorincz-Comi, Yihe Yang, Gen Li, and Xiaofeng Zhu. Mrbee: A novel bias-corrected multivariable mendelian randomization method. *biorxiv, 523480*, 2023.

[21] Noah Lorincz-Comi, Yihe Yang, Gen Li, and Xiaofeng Zhu. Mrbee: A novel bias-corrected multivariable mendelian randomization method. *bioRxiv*, pages 2023–01, 2023.

[22] Rahul Mazumder, Trevor Hastie, and Robert Tibshirani. Spectral regularization algorithms for learning large incomplete matrices. *The Journal of Machine Learning Research*, 11:2287–2322, 2010.

[23] Paul J Newcombe, David V Conti, and Sylvia Richardson. Jam: a scalable bayesian framework for joint analysis of marginal snp effects. *Genetic*

*epidemiology*, 40(3):188–201, 2016.
